# Multiple sclerosis disease-modifying therapies and COVID-19 vaccines: A practical review and meta-analysis

**DOI:** 10.1101/2022.02.12.22270883

**Authors:** Masoud Etemadifar, Hosein Nouri, Maristella Pitzalis, Maria Laura Idda, Mehri Salari, Mahshid Baratian, Sepide Mahdavi, Amir Parsa Abhari, Nahad Sedaghat

## Abstract

**Importance:** An evidence-based appraisal of the COVID-19 vaccination policies among people with multiple sclerosis (pwMS) with respect to disease-modifying therapies (DMT) is important for our understandings and their further management.

**Objective:** To synthesize the available evidence concerning the effect of DMTs on COVID-19 vaccination immunogenicity and effectiveness.

**Data Sources:** We searched MEDLINE, Scopus, Web of Science, MedRxiv, and Google Scholar from January 2021 until January 2022.

**Study Selection:** The exclusion criteria included: not a primary investigation; retracted/withdrawn; no eligible participants – people with no history/evidence of previous COVID-19 and corticosteroid administration within two months of vaccination; no eligible exposures – all nine DMT classes; and no eligible comparators – DMT-unexposed at the time of vaccination.

**Data Extraction and Synthesis:** Entries were assessed independently by two reviewers for eligibility and quality. Dichotomized data was extracted by two reviewers in accordance with Cochrane guidelines, and were pooled using either Peto fixed-effects or Inverse-variance random-effects methods.

**Main Outcomes and Measures:** Main outcomes were i) B-cell response, measured by seroconversion odds ratio (OR); ii) T-cell response, measured by interferon-gamma release response OR, and CD4+/CD8+ activation-induced marker+ OR. Further outcomes including immunity waning speed and breakthrough COVID-19 incidence/severity were synthesized narratively.

**Results:** Data from 28 studies (5,025 pwMS and 1,635 healthy participants) after COVID-19 vaccination suggests mildly-lower B-cell responses in teriflunomide- and alemtuzumab-treated, extensively-lower B-cell responses in sphingosine-1-phosphate receptor modulator (S1PRM)- and anti-CD20 (aCD20)-treated, and lower T-cell responses in interferon-, S1PRM-, alemtuzumab- and cladribine-treated pwMS. Every ten-week increase in aCD20-to-vaccine period is associated with a 1.94-time (95%CI: 1.57, 2.41, P<0.00001) increase in odds of seroconversion. B-cell-depleting therapies seem to accelerate post-vaccination humoral waning, and booster immunogenicity is predictable with the same factors affecting the priming vaccination. Furthermore, comparatively-increased breakthrough COVID-19 incidence and severity is being observed only among S1PRM- and anti-CD20-treated pwMS – i.e., among the pwMS with extensively-blunted B-cell response, despite adequate T-cell responses in the aCD20-treated. To date, pwMS on only-T-cell-blunting DMTs have not shown increased susceptibility to breakthrough COVID-19.

**Conclusion and Relevance:** The implemented vaccination strategy to date has been effective for pwMS on all DMTs other than S1PRM and aCD20. As B-cell immunity seems to be a more important predictor of vaccine effectiveness than T-cell immunity, optimization of humoral immunogenicity and ensuring its durability among pwMS on DMTs are the necessities of an effective COVID-19 vaccination policy.

## 1. Introduction

From the beginning of the coronavirus disease 2019 (COVID-19), global mass vaccination has been the most prominent effort of humanity to end the reign of severe acute respiratory syndrome coronavirus 2 (SARS-CoV-2). Ever since, many vaccines have been developed, all with reasonable safety and efficacy profiles (more information available at: https://covid19.trackvaccines.org/agency/who).

As the effectiveness of COVID-19 vaccination was thought to be altered in people with multiple sclerosis (pwMS) who receive immunomodulatory disease-modifying therapies (DMTs), expert panels issued adjusted vaccination guidelines for pwMS based on previous knowledge of the DMTs’ mechanisms of action and preliminary real-world evidence ^1-4^. These guidelines mostly concerned people on sphingosine 1-phosphate receptor modulators (S1PRM), anti-CD20 therapies (aCD20), and other B-cell depleting therapies (BCDT), as they were thought to blunt COVID-19 vaccines’ immunogenicity.

Furthermore, administration of booster – in most cases, third – doses of COVID-19 vaccines was recommended after observation of waning humoral immunity ^5^ and clinical effectiveness ^6-8^. It was stressed after showing to be effective against the vaccine-escape ^9^ Omicron variant of the SARS-CoV-2 ^10-12^ – regardless of the priming regimen ^13,14^. Among the pwMS, homologous booster doses were tested as a strategy to immunize the ones who did not seroconvert following their priming regimen.

Now, several months after the mass vaccination of pwMS around the globe, the available real-world evidence seems adequate for a practical appraisal of the COVID-19 vaccination policies among pwMS with respect to their DMT. Hence, in this systematic review and meta-analysis study, we aimed to gather and synthesize the available evidence and highlight the gaps in the literature, providing a direction guide for future research, facilitating further policy makings, and enabling evidence-based management of pwMS.

We hereby reported and discussed the results of our study (PROSPERO id: CRD42021278107) in accordance with the Preferred Reporting Items for Systematic Reviews and Meta-Analyses (PRISMA) statement (available from: http://www.prisma-statement.org). The detailed methods of our study could be accessed from the online Supplementary Material.

## 2. Results and Discussion

Overall, 28 studies including 5,025 pwMS and 1,635 healthy controls were synthesized (***Fig. 1, Table 1***). One study ^15^ was excluded despite containing eligible participants because their data could not be extracted. The studies differed in outcome measurement methods, settings, number of participants, and the administered vaccines. The assessed vaccines used either mRNA (BNT162b2, mRNA-1273), adenoviral vector (AV) (Ad26.COV2.S, ChAdOx1), or inactivated (CoronaVac, BBIBP-CorV) platforms. Other prominent points of heterogeneity were the used assays, the number/types of assessed DMTs, the usage of different comparators, e.g., healthy participants, pwMS on no DMT etc., and the timepoints of obtaining samples from participants.

**Table 1.**
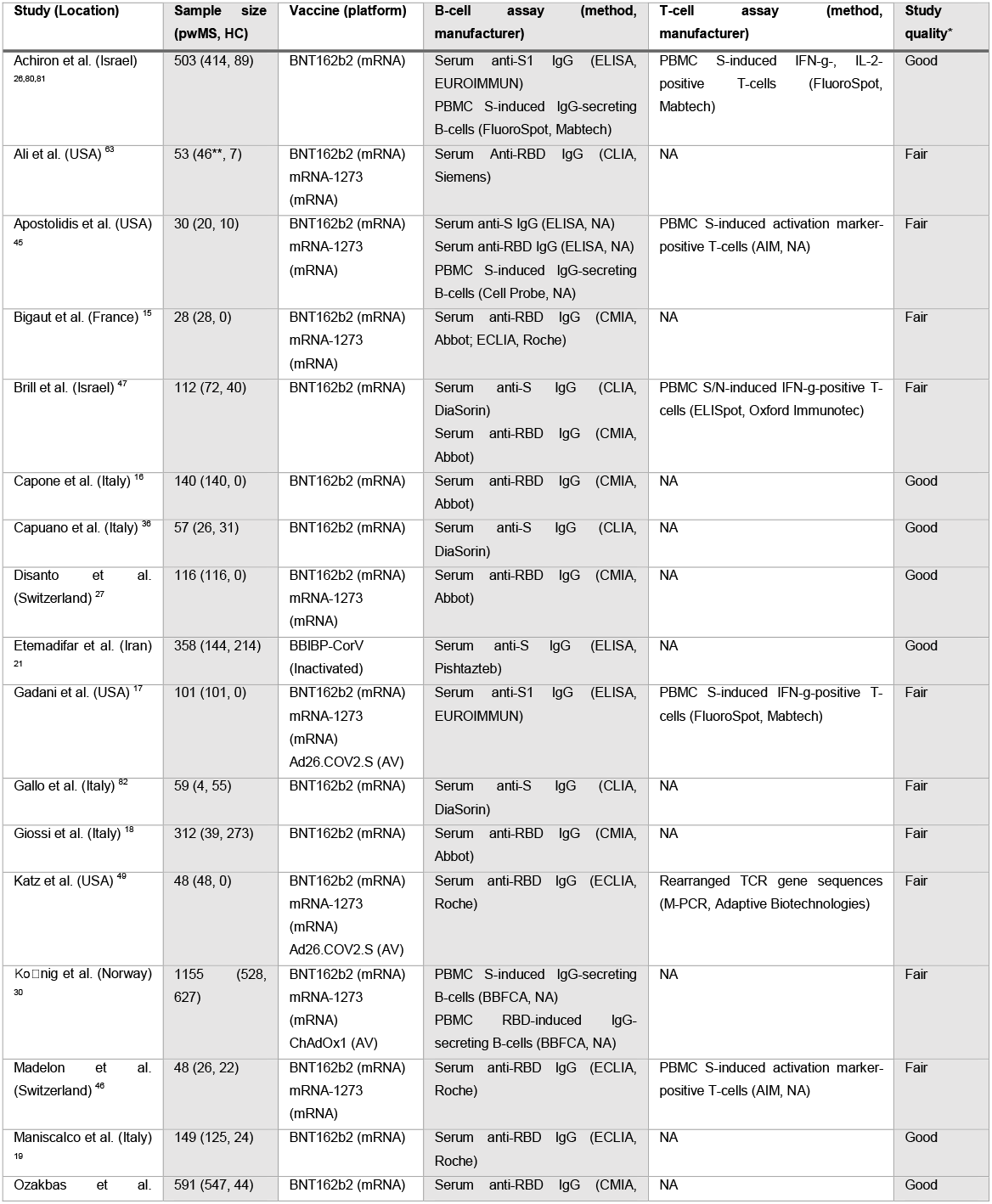
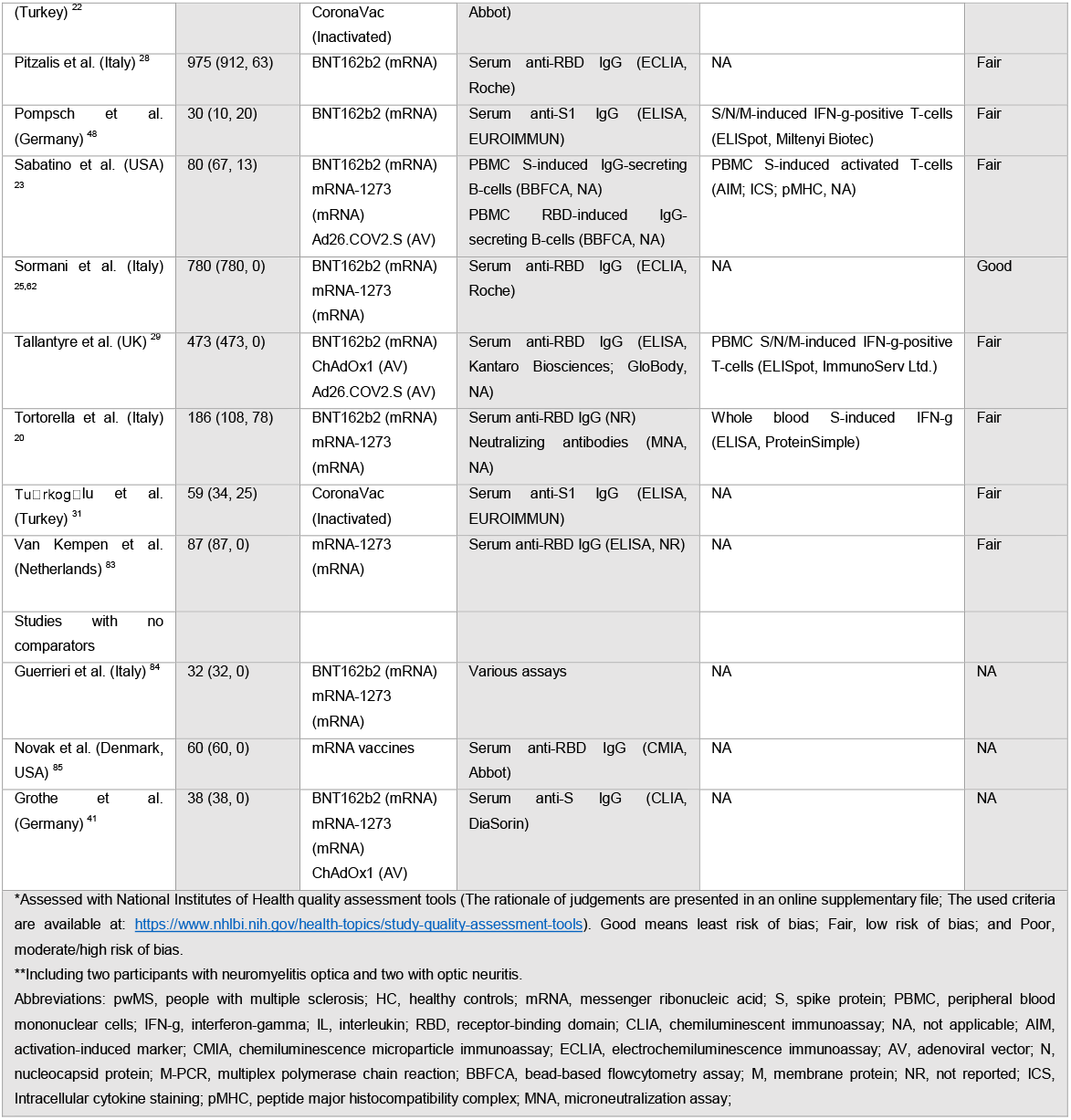
Characteristics of included studies.

**Fig. 1.**
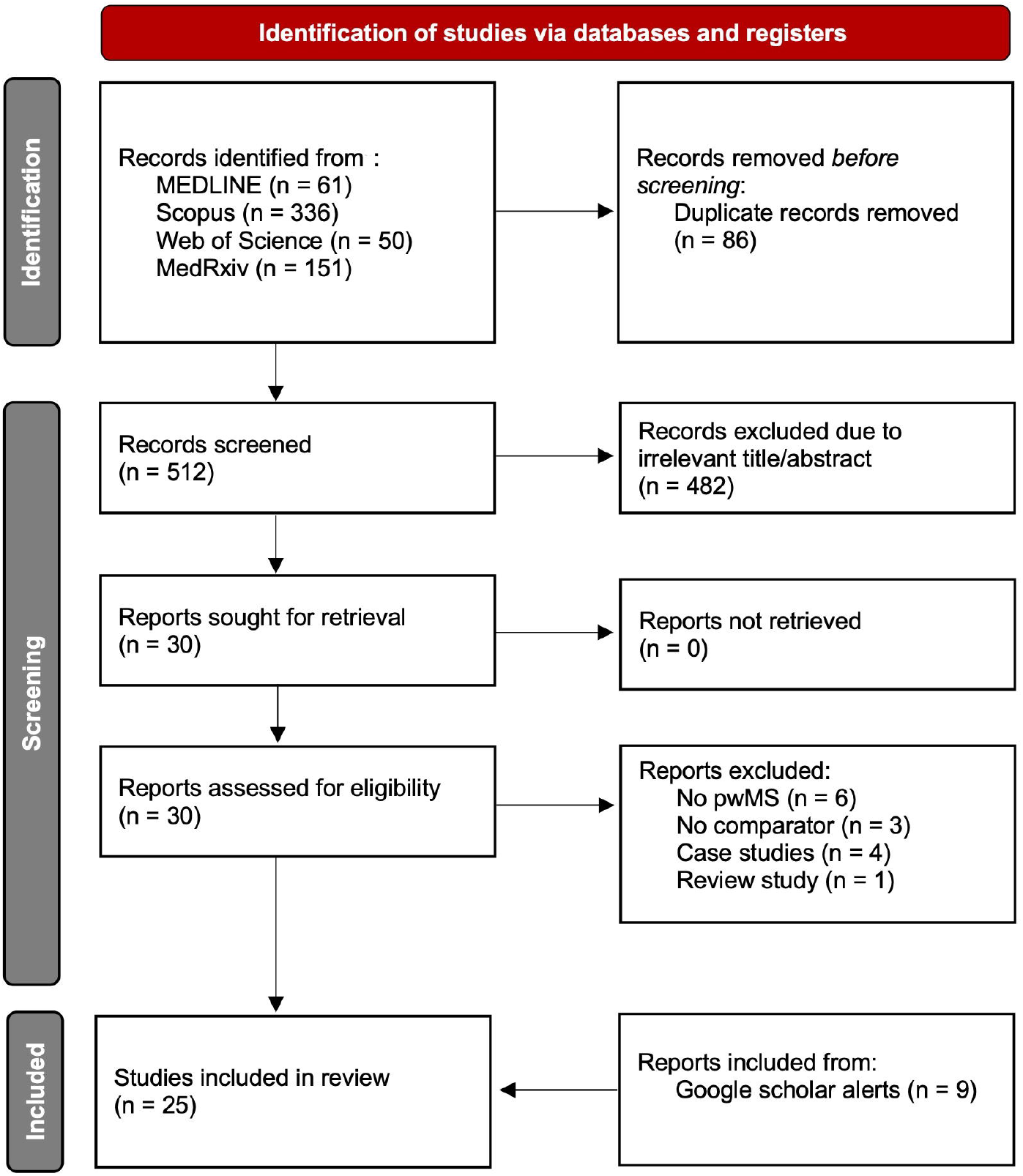
PRISMA Flow Diagram. Abbreviation: pwMS, people with multiple sclerosis.

The results of individual studies, heterogeneity tests, forest and funnel plots, and the detailed rationale behind each quality assessment – based on NIH tools – is accessible from the online Supplementary Material. The pooled measures are summarized in both ***Table 2 and Figure 2***, along with their certainty based on GRADE.

**Table 2.** Summary of Findings. Results with statistically-significant P-values are bolded.

| Factors (ref) | Post-vac outcomes |  |  |  |  |  |  |  |
| --- | --- | --- | --- | --- | --- | --- | --- | --- |
|  | B-cell response |  | T-cell response |  |  |  |  |  |
|  | Seroconversion |  | IFN-g release |  | CD4+ AIM+ |  | CD8+ AIM+ |  |
|  | OR<br>(95% CI) | P-value<br>(certainty*) | OR<br>(95% CI) | P-value<br>(certainty*) | OR<br>(95% CI) | P-value<br>(certainty*) | OR<br>(95% CI) | P-value<br>(certainty*) |
| DMT (UX) |  |  |  |  |  |  |  |  |
| -IFN | 0.84<br>(0.38, 1.83) | 0.66<br>Moderate<sup>1,2</sup> | 0.02<br>(0.00, 0.14) | <0.01<br>Very Low<sup>2,4,5</sup> | Ni |  | Ni |  |
| -GA | 0.87<br>(0.31, 2.42) | 0.79<br>Moderate<sup>1,2</sup> | Ni |  | NE |  | 0.62<br>(0.04, 9.00) | 0.72<br>Very Low<sup>1,2,4,5</sup> |
| -DMF | 1.98<br>(0.96, 4.09) | 0.07<br>Moderate<sup>1,2</sup> | Ni |  | NE |  | 3.78<br>(0.18, 78.38) | 0.39<br>Very Low<sup>1,2,4,5</sup> |
| -TERI | 0.38<br>(0.16, 0.90) | 0.03<br>Very Low<sup>2,3</sup> | Ni |  | Ni |  | Ni |  |
| -S1PRM | 0.04<br>(0.03, 0.06) | <0.00001<br>High<sup>1</sup> | 0.04<br>(0.02, 0.07) | <0.00001<br>Low<sup>1,2,3</sup> | 0.01<br>(0.00, 0.18) | 0.001<br>Very Low<sup>2,4,5</sup> | 0.95<br>(0.08, 10.71) | 0.97<br>Very Low<sup>1,2,4,5</sup> |
| -NTZ | 0.53<br>(0.24, 1.18) | 0.12<br>Low<sup>1,2,4</sup> | 1.00<br>(0.20, 5.12) | 1.00<br>Very Low<sup>1,2,4,5</sup> | NE |  | 3.95<br>(0.23, 69.44) | 0.35<br>Very Low<sup>1,2,4,5</sup> |
| -CLAD | 0.41<br>(0.15, 1.11) | 0.08<br>Low<sup>1,2,5</sup> | 0.01<br>(0.00, 0.04) | <0.00001<br>Low<sup>1,2,5</sup> | Ni |  | Ni |  |
| -ALEM | 0.32<br>(0.10, 0.96) | 0.04<br>Very Low<sup>2,3</sup> | Ni |  | Ni |  | Ni |  |
| -aCD20 | 0.05<br>(0.04, 0.06) | <0.00001<br>High<sup>1</sup> | 1.12<br>(0.62, 2.05) | 0.70<br>Moderate<sup>1,3</sup> | 1.13<br>(0.17, 7.61) | 0.90<br>Moderate<sup>1,2</sup> | 2.54<br>(0.89, 7.27) | 0.08<br>Moderate<sup>1,5</sup> |
| aCD20 infusion-to-vac<br>(per 10 weeks) | 1.94<br>(1.57, 2.41) | <0.00001<br>High<sup>1</sup> | Ni |  | Ni |  | Ni |  |
| *Based on GRADE approach (Baseline: moderate due to observational nature of studies; ▲upgrade; ▼downgrade): |  |  |  |  |  |  |  |  |
| 1. ▲ Result in line with quantitative analyses in individual studies; 2. ▼ Low count of studies with measurable relative effect; 3. ▼ Inconsistency; 4. ▼ Risk of missing results; and 5. ▼ Imprecision. |  |  |  |  |  |  |  |  |
| Abbreviations: ref, reference; vac, vaccination; IFN-g, interferon-gamma; AIM, activation-induced marker; OR, odds ratio; CI, confidence interval; DMT, disease-modifying therapy; UX, unexposed; IFN, interferons; Ni, no information; GA, glatiramer acetate; NE, not estimable; TERI, teriflunomide; S1PRM, sphingosine-1-phosphate receptor modulators; NTZ, natalizumab; CLAD, cladribine; ALEM, alemtuzumab; aCD20, anti-CD20 therapies. |  |  |  |  |  |  |  |  |

**Fig. 2.**
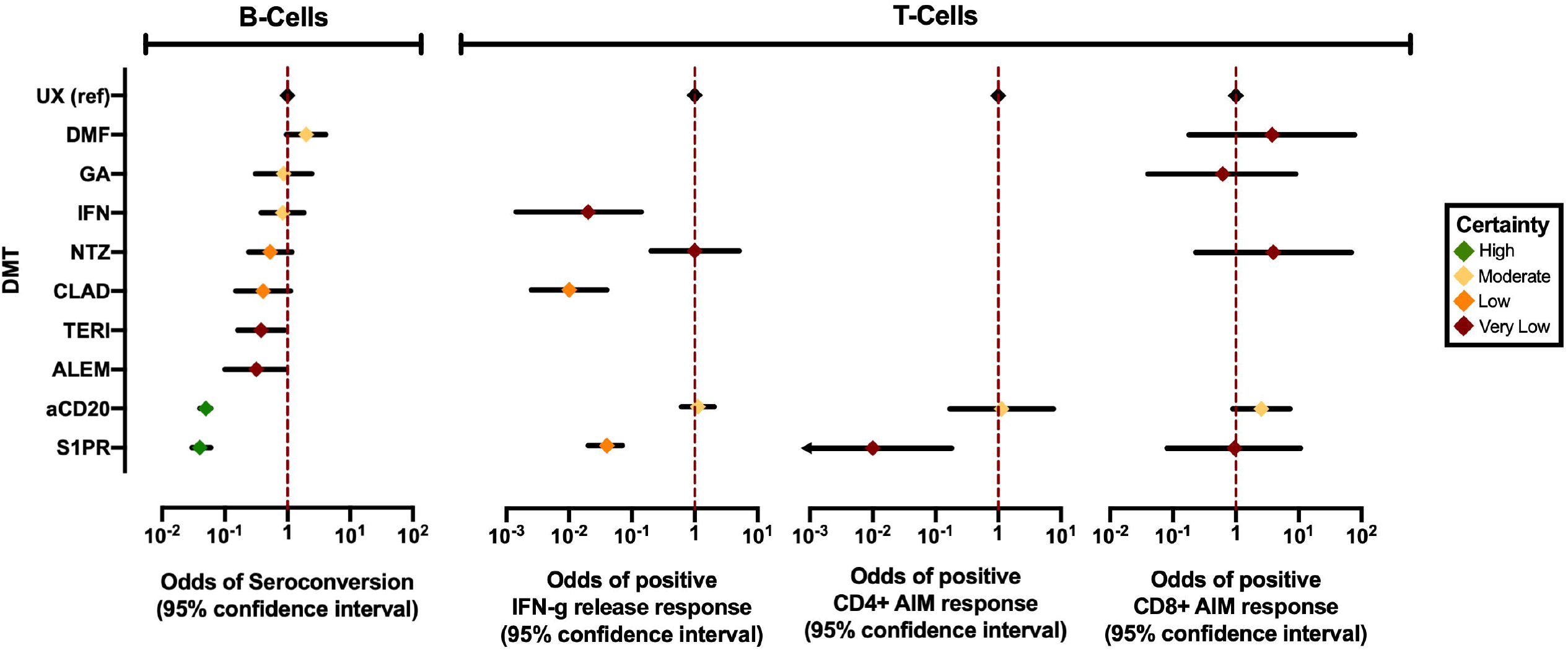
Summary forest plot of the pooled results. Abbreviations: UX, unexposed; ref, reference; DMF, dimethyl fumarate; GA, glatiramer acetate; IFN, interferons; NTZ, natalizumab; CLAD, cladribine; TERI, teriflunomide; ALEM, alemtuzumab; aCD20, anti-CD20; S1PR, sphingosine-1-phosphate receptor modulators; IFN-g, interferon-gamma; AIM, activation-induced marker.

### 2.1. Effect of DMTs on COVID-19 vaccines’ immunogenicity

#### 2.1.1. Interferons (IFN)

Moderate-certainty evidence does not suggest decreased odds of post-vaccination seroconversion in the pwMS on IFNs compared to people unexposed (UX) to DMTs (OR [95%CI]: 0.84 [0.38, 1.83], P=0.66) (Supplementary Figure 1). No seronegative pwMS on IFN were present in four studies ^16-19^. Quantitative analysis in studies also did not suggest lower concentrations of antibodies among these pwMS post-vaccination. In one study, significantly higher concentrations of anti-Spike (S) receptor binding domain (RBD) IgG were detected among pwMS on IFNs, compared to healthy controls ^19^. Although the authors suggested that IFN-beta 1a therapy may promote the post-vaccination antibody responses in pwMS, this finding was not observed in other studies.

Very-low-certainty evidence from one study ^20^ showed lower extents of interferon-gamma release response to the S antigen in samples from pwMS on IFNs, suggesting blunted T-cell response among these people compared to healthy controls (OR [95%CI]: 0.02 [0.00, 0.28], P<0.01). Both CD4+ and CD8+ T-cell responses were reduced in samples of pwMS on IFN compared to UX people, according to flow cytometric analysis in Tortorella et al. study ^20^.

#### 2.1.2. Glatiramer Acetate (GA)

Moderate-certainty evidence did not suggest decreased odds of post-vaccination seroconversion in the pwMS on GA compared to the UX people (OR [95%CI]: 0.87 [0.31, 2.42], P=0.79) (Supplementary Figure 2). Post-vaccination seronegative pwMS on GA were only present in two studies ^21,22^ – all being among the ones receiving inactivated vaccination. Quantitative analysis in suggested no difference in post-vaccination antibody concentrations between GA-treated pwMS and the UX people in any study.

One study with limited sample size utilizing AIM assays ^23^ (very low-certainty evidence), suggested decreased odds of positive response neither in CD4+ (OR not measurable) nor CD8+ (OR [95%CI]: 0.62 [0.04, 9.00], P=0.72) T-cells. Furthermore, it is worth mentioning that adequate interferon-gamma release responses were present after SARS-CoV-2 *infection* in these people ^24^, suggesting favorable T-cell responses.

#### 2.1.3. Dimethyl Fumarate (DMF)

Moderate-certainty evidence did not suggest any decrease in odds of post-vaccination seroconversion among pwMS on DMF compared to UX people (OR [95%CI]: 1.98 [0.96, 4.09], P=0.07) (Supplementary Figure 3). Seven studies ^16,18,19,22,23,25,26^ did not contain any seronegative pwMS on DMF following mRNA or inactivated vaccination; Quantitative analysis suggested no difference in post-vaccination antibody concentrations compared to UX people.

Similar to GA-treated pwMS, evidence on T-cell responses among these pwMS was limited to one study with a limited sample size ^23^ (very-low-certainty evidence), which by utilizing AIM assays, suggested no decrease in odds of positive responses in subsets of CD4+ (OR not measurable) and CD8+ (OR [95%CI]: 3.78 [0.18, 78.38], P=0.39) T-cells. Similarly, interferon-gamma release responses were sufficient in DMF-treated pwMS after SARS-CoV-2 *infection*, suggesting adequate T-cell response ^24^.

#### 2.1.4. Teriflunomide (TERI)

Inadequate number of studies with considerable heterogeneity (very-low-certainty evidence) suggest decreased odds of post-vaccination seroconversion in pwMS on TERI compared to UX people (OR [95%CI]: 0.38 [0.16, 0.90], P=0.03) (Supplementary Figure 4). Due to low number of mRNA vaccine studies with measurable relative effect and therefore, the uncertainty of their pooled measures, the difference between the inactivated and mRNA measures did not reach statistical significance (Chi^2^=2.91, P=0.09). Nevertheless, considering that none of the TERI-treated pwMS in any of the studies ^16,18,22,25-28^ remained seronegative after mRNA vaccination, the suggested blunt caused by TERI may not be generalizable to the mRNA-vaccinated pwMS. Furthermore, lower antibody concentrations compared to UX people were observed in pwMS on TERI homogenously in all studies regardless of the used vaccine, but reached statistical significance only in one ^28^. TERI’s mechanism of action – which involves inhibition of rapidly-dividing cells, including activated B-cells – may explain this observation.

Furthermore, no evidence was found regarding the T-cell responses in these pwMS following vaccination; Assessing the vaccine-induced T-cell responses in TERI-treated pwMS is therefore encouraged.

#### 2.1.5. Sphingosine-1-phosphate receptor modulators (S1PRM)

High-certainty evidence confirms significantly lower odds of post-vaccination seroconversion in pwMS on S1PRM compared with UX people (OR [95%CI]: 0.04 [0.03, 0.06], P<0.00001) (Supplementary Figure 5). All included studies ^16,17,19-22,25-31^ found significantly lower concentrations of antibodies following vaccination in these pwMS compared to UX people. Studies with heterogenous effect measures (moderate-certainty evidence) indicate that with the current vaccination strategy, pwMS on S1PRM are 25 times (95%CI: 16.66, 33.33) less likely to show anti-S1, and 8.33 times (95%CI: 3.70, 20) less likely to show anti-S seroconversion following COVID-19 vaccination (Chi^2^=7.24, P<0.01). Paradoxical to the healthy population, low-certainty evidence – due to limited count of inactivated vaccine studies – suggests that among the pwMS on S1PRM, odds of anti-S1 seroconversion is higher with inactivated vaccines compared to mRNA and AV vaccines (Chi^2^=11.97, P<0.001). Although theoretically reasonable ^32^, head-to-head mRNA-AV ^29^ and mRNA-inactivated ^22^ immunogenicity comparisons in S1PRM-treated pwMS have suggested the opposite. Hence, the need for more replication of inactivated/mRNA/AV comparisons is clearly sensed for pwMS on S1PRM.

Furthermore, interferon-gamma release assays in two studies ^20,26^ (low-certainty evidence) suggested decreased odds of positive T-cell response in pwMS on S1PRM (OR [95%CI]: 0.04 [0.02, 0.07], P<0.00001) (Supplementary Figure 6). AIM assay in another study ^23^ (very-low-certainty evidence) did not suggest decreased odds of CD8+ T-cell response in these pwMS (OR [95%CI]: 0.95 [0.08, 10.71], P=0.97) but suggested decreased odds of CD4+ T-cell responses (OR [95%CI]: 0.01 [0.00, 0.18], P=0.001) compared to UX people.

Additionally, among the pwMS on S1PRM who failed to seroconvert following priming vaccination, one study showed that administration of booster doses increased anti-S1 antibody concentrations, but promoted seroconversion only in 2/29 (7%) ^33^.

The trafficking inhibition of lymphocytes, their restriction to lymphatics, and hence, the peripheral lymphopenia seen in pwMS on S1PRM explains the lower T-cell reactivity observed in peripheral blood samples – not to mention S1PRM’s documented inhibitive effect on T-cell activation ^34^. The former reason may also explain the blunted humoral responses, as the one-way flow of lymph from peripheral to central areas restricts trafficking of lymphocytes to peripheral areas, inhibiting proper exposure of lymph-trapped lymphocytes to immunizing materials of the vaccines which are administered peripherally/locally. As systemic SARS-CoV-2 infection mounts adequate immunization among pwMS on S1PRM ^32,35^, it can be hypothesized that immunogenicity in these people is subject to wider (e.g., systemic instead of local/peripheral) exposure to immunogens. When the lymphocytes’ ability to reach the immunogens is inhibited, the immunogens should reach the lymphocytes themselves or immunization will not develop.

#### 2.1.6. Natalizumab (NTZ)

Compared to UX people, low-certainty evidence did not confirm lower odds of anti-S1 seroconversion among pwMS on NTZ following vaccination (OR [95%CI]: 0.53 [0.24, 1.18]) (Supplementary Figure 7). All pwMS on NTZ in seven studies – including the only two studies using anti-S assays with extractable data ^23,36^ – seroconverted following vaccination ^16,17,23,25,26,28,36^. Although relative effect was measurable in only two of them which contained seronegative UX people ^26,28^. Unlike all the other studies, pwMS on NTZ in one study ^28^ showed significantly lower post-vaccination antibodies compared to UX people. Interferon-gamma release ^17^ and AIM ^23^ assays (very low-certainty evidence) did not suggest blunted post-vaccination T-cell responses in pwMS on NTZ compared to UX people.

NTZ, an anti-α4-integrin monoclonal antibody, implements its effect by inhibiting lymphocyte extravasation; however, unlike S1PRM, its lymphocyte trafficking inhibition does not trap the lymphocytes in the lymphatic system – i.e., it does not cause peripheral lymphopenia. Although their trafficking abilities are inhibited, the preserved presence of lymphocytes in blood flow – which, unlike the lymph flow, can be from central to peripheral areas as well – may be the reason NTZ does not blunt vaccination-induced immunization as much as S1PRM.

#### 2.1.7. Cladribine (CLAD)

Pooled low-certainty evidence (Supplementary Figure 8) confirms no difference in odds of anti-S1 seroconversion among pwMS on CLAD compared to UX people (OR [95%CI]: 0.41 [0.15, 1.11], P=0.08). No evidence was found regarding anti-S seroconversion. In five studies ^16,19,20,22,25^, all CLAD-treated pwMS seroconverted following vaccination similar to the UX people, preventing relative effect measurement. Quantitative analysis in no studies suggested lower concentrations of post-vaccination antibodies among them. Assessment of vaccination-induced T-cell response was limited to one study ^20^ (very-low-certainty evidence); it showed lower odds of positive S-induced interferon-gamma release responses in samples from pwMS on CLAD compared to UX people (OR [95%CI]: 0.01 [0.00, 0.04], P<0.00001).

Compared to its effect on the T-cell lineage, CLAD’s effect on the B-cells is more extensive but less durable ^37-40^. As interpreted, this has been translated into observation of proper humoral despite blunted cellular immunization following COVID-19 vaccination among pwMS on CLAD. Furthermore, the time since the last CLAD dose theoretically affects humoral responses; This was suggested especially by Achiron et al. study ^26^ but did not reach statistical significance, and was not confirmed by other studies ^25,28,41^; It seems the implemented guidelines ^1-4^ have suggested an adequate amount of post-CLAD vaccination delay to prevent blunted humoral responses, and therefore, made the probable effect unmeasurable. Additionally, although CLAD depletes the memory B-cells ^42^, a preliminary study suggested its subsequent doses will not alter pre-existing humoral memory ^43^; Still, there is limited evidence that the longevity of COVID-19 vaccine-induced humoral responses is lower in pwMS on CLAD ^26^. Replicative studies measuring the immunity waning speed in these pwMS after COVID-19 vaccination are, therefore, required to determine whether they require personalized booster schedules.

#### 2.1.8. Alemtuzumab (ALEM)

Pooled very-low-certainty evidence (Supplementary Figure 9) suggests lower odds of anti-S1 seroconversion among pwMS on ALEM compared to UX people (OR [95%CI]: 0.32 [0.10, 0.96], P=0.04). One study assessed anti-S seroconversion, but its data could not be extracted ^30^. PwMS on ALEM in three studies ^16,25,28^ showed 100% seroconversion rates similar to the UX people in two of them ^16,25^, and none of the studies suggested lower concentrations of post-vaccination antibodies among them – not even the study that indicated lower odds of seroconversion ^26^. Vaccination-induced T-cell responses were not assessed in any of the included studies, still, ALEM’s durable effect of T-cell lineage ^44^ suggests that they are blunted .

ALEM, an anti-CD52 monoclonal antibody, is known to significantly deplete B- and T-cells shortly after administration. ALEM’s short-term effect on B- and T-cell dynamics is relatively similar to CLAD ^40,44^. Hence, although the time from the last ALEM infusion affects seroconversion ^26^, this effect is currently not measurable as the implemented guidelines ^1-4^ seem to have suggested an adequate amount of delay. Similar to other BCDT, further studies measuring comparative immunity waning speeds in pwMS on ALEM are needed to determine whether they require more frequent boosters.

#### 2.1.9. Anti-CD20 therapies (aCD20)

High-certainty evidence confirms lower odds of seroconversion following COVID-19 vaccination among pwMS on aCD20 compared to UX people (OR [95%CI]: 0.05 [0.04, 0.06], P<0.00001) (Supplementary Figure 10). Studies with heterogenous effect measures (moderate-certainty evidence) indicate that with the currently-implemented strategies, pwMS on aCD20 are 20 times (95%CI: 16.66, 25) less likely to seroconvert for anti-S1, and 12.5 times (95%CI: 7.69, 20) less likely to seroconvert for anti-S antibodies (Chi^2^=2.76, P=0.10) following COVID-19 vaccination. Quantitative analyses in all included studies confirmed this observation. Furthermore, evidence indicates with high certainty that every 10-week delay in subsequent aCD20 infusion is associated with a 1.94-time (95%CI: 1.57, 2.41, P<0.00001) increase in seroconversion odds of pwMS on aCD20 (Supplementary Figure n).

Regarding the T-cell responses, compared to UX people, moderate-certainty evidence does not suggest different odds of positive post-vaccination T-cell interferon-gamma release responses (OR [95%CI]: 1.12 [0.62, 2.05], P=0.70) (Supplementary Figure 12), CD8+ (OR [95%CI]: 2.54 [0.89, 7.27], P=0.08) (Supplementary Figure 13), and CD4+ (OR [95%CI]: 1.13 [0.17, 7.61], P=0.90) T-cell AIM responses. Quantitative analyses in most studies ^17,20,23,29,45-48^ were in line with the dichotomized evidence. Multiplex polymerase chain reaction assay in one study ^49^ indicated positive adaptive T-cell responses among 100% of seronegative pwMS on aCD20 following vaccination.

Furthermore, the preliminary evidence indicates significant decline in seropositivity rates of pwMS on aCD20 six months after their second dose ^50,51^. Homologous mRNA boosters in pwMS on aCD20 promoted T-cell responses ^52^, while humoral responses were still heavily dependent on the serostatus following the priming regimen, and B-cell dynamics at the time of booster administration ^33,51-53^; In other words, the booster doses did not promote humoral immunization in pwMS on aCD20 who did not seroconvert following priming vaccination, unless their B-cells were reconstituted. Studies among people on aCD20 with diseases other than MS ^54,55^ support the same conclusion.

Similar to pwMS on other DMTs, the COVID-19 vaccines’ immunogenicity among pwMS on aCD20 could be considered a translation of the previously-determined B- and T-cell dynamics in them ^40,56^, based on which the current guidelines recommended a 12-to-36-week window between aCD20 infusion and COVID-19 vaccination ^1-4^. However, the presented evidence suggests that the mentioned interval, although increases the odds, will not be adequate to reverse the humoral blunts in pwMS on aCD20 (Fig. 3). The alterations in the dynamics of B-cells in people receiving aCD20 last for years according to the unpublished results from the NCT00676715 phase-II extension trial ^57^, suggesting durable, long-lasting benefits of aCD20 without subsequent dosing ^58^. However, this durable effect of aCD20 has shown to be able to alter vaccine immunogenicity for as long as three years, as observed in people with hematological malignancies ^59^. Hence, prior B-cell profiling and post-vaccination serological screening may be the necessities of an effective personalized vaccination strategy in pwMS who received aCD20 at any time point within three years.

**Fig. 3.**
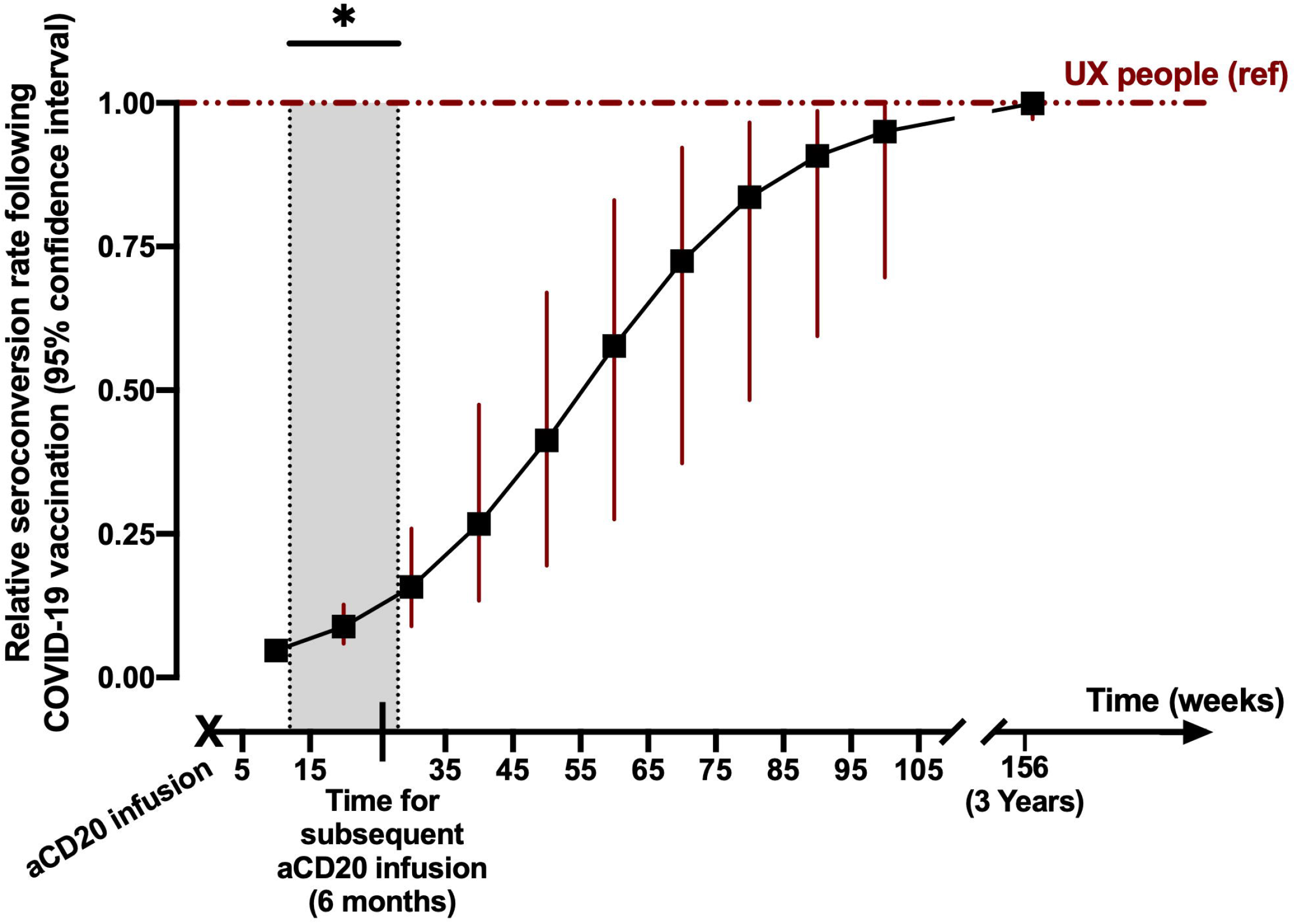
Schematic curve showing the association of post-vaccination seroconversion rates with time since last anti-CD20 infusion. *Current guideline recommendations on minimum delay of vaccination after anti-CD20 infusion. Abbreviations: UX, unexposed; aCD20, anti-CD20.

### 2.2. Vaccine types

Based on phase-III data, the efficacy profiles of different available COVID-19 vaccine types (i.e., mRNA, AV, inactivated, and protein-based) seems to correlate to their anti-S/S1 humoral immunogenicity – both in healthy people ^60,61^ and pwMS ^62^. Head-to-head comparisons of the available COVID-19 vaccines’ humoral immunogenicity among pwMS reveals the superiority of mRNA-1237 over BNT162b2 (both mRNA-based) ^20,25,63^ – probably due to higher concentrations of active material, BNT162b2 and mRNA-1237 (mRNA) over ChAdOx1 and Ad26.COV2 (AV) ^17,23,29,49^ and BNT162b2 (mRNA) over CoronaVac (inactivated) ^22^ – although humoral immunization did not differ significantly in pwMS on aCD20 receiving BNT162b2 and CoronaVac in Ozakbas et al. ^22^ study. The choice of a specific vaccine type among pwMS is encouraged and should be based on an individualized risk/benefit assessment with careful consideration of their COVID-19 risk factor profile ^64,65^, their DMT, and the availability/cost-effectiveness of the vaccine ^66,67^.

### 2.3. Moving into the clinic

While the serum anti-S/S1 assays are deemed predictive of its neutralizing activity ^68,69^, and the serum neutralizing activity predictive of the clinical protection against symptomatic SARS-CoV-2 infection ^70^, pwMS with adequate humoral responses – mostly on DMTs other than aCD20 and S1PRM – would theoretically show adequate protection against SARS-CoV-2. However, in the absence of humoral immunization, it is doubted if the T-cells could provide adequate clinical protection among pwMS on aCD20 and S1PRM. The current real-world evidence confirms the predictive effect of seroconversion on post-vaccination COVID-19 incidence and severity ^62^, and indicates rising comparative incidence and severity of COVID-19 among pwMS on S1PRM and aCD20 following vaccination of pwMS ^62,71-73^. The less-extensive humoral blunts in pwMS on TERI – and possibly ALEM – do not seem to have had any significant effect on vaccine effectiveness. Regarding the protective effect of T-cell responses, currently, the only clue lies within the Etemadifar et al. ^72^ study – although not confirmed by a larger study ^73^; it showed that among the vaccinated pwMS, the ones on aCD20 experienced lower incidence and severity of COVID-19 in comparison with those on fingolimod. The former are known to be prone to worse COVID-19 outcomes ^64^ but show robust post-vaccination T-cell responses, while the latter do not develop proper T-cell immunization following vaccination. Still, the practical protective effect of T-cell responses in the absence of antibodies could neither be confirmed nor measured until further real-world evidence becomes available.

## 3. Conclusion

The present analysis highlight and corroborate the relevance for an optimal treatment strategy in pwMS before COVID-19 vaccination. It was demonstrated that the current vaccination strategy has failed to promote adequate humoral immunity in aCD20-and S1PRM-treated pwMS, which is being translated into low clinical effectiveness of COVID-19 vaccines among them – despite adequate T-cell responses in the ones on aCD20. Their susceptibility to worse COVID-19 outcomes, and the dependency of COVID-19 vaccines’ humoral immunogenicity to the B-cell dynamics at the time of administration – and therefore, the timing of aCD20 infusion – stress the importance of personalizing vaccination strategies for pwMS on aCD20 with respect to their B-cell profiles and aCD20 infusion timings. Theoretically and based on limited evidence, mode of action and administration method may be important factors to consider also when vaccinating S1PR modulator-treated pwMS, while more evidence is needed to support this claim. Milder humoral and considerable T-cell response blunts – also depending on dosage timings, and higher immunity waning speeds may be present in pwMS on CLAD and ALEM, which subject to confirmation by further evidence, stresses the importance of earlier booster administrations among them. TERI may also cause a humoral immunogenicity blunt, however, being less extensive and clinically-irrelevant based on current evidence; PwMS on TERI may not require countering policies other than being provided with reliable information about the importance of booster doses. Evidence to date does not indicate any significant effect of IFN, GA, DMF, and NTZ on COVID-19 vaccines’ immunogenicity and effectiveness.

Additionally, as heterologous boosters among healthy people showed to be more immunogenic and effective ^74^, further replication among pwMS with heterologous regimens is encouraged, especially among the ones primed with inactivated vaccines – as samples from inactivated-vaccinated people show less neutralizing activity against the Omicron variant ^75^, and the heterologous boosters with superiority of mRNA ^13,14,76-78^ over AV ^79^ have shown to be more immunogenic than homologous boosters in healthy people receiving inactivated priming regimens.

## Supporting information

Quality Assessment

## Data Availability

Not applicable. This work is a secondary study and no data was produced.

## 4. Acknowledgements

We would like to thank Prof. Mohammad Reza Maracy for his valuable methodological consultations.

