## Supplementary material for "Multiple sclerosis disease-modifying therapies and COVID-19 vaccines: A practical review and meta-analysis": Methods and Supplementary Figures

### 1. Methods

We hereby report our systematic review and meta-analysis study methods in accordance with the Preferred Reporting Items for Systematic Reviews and Meta-Analyses (PRISMA) statement (available from: <http://www.prisma-statement.org>).

#### 1.1. The Search

Based on the objectives of the review, a comprehensive search of the MEDLINE, Scopus, and Web of Science was performed with consideration of their specific vocabulary and indexing approaches. To consider the unpublished data, the medRxiv preprint server and Google Scholar were also searched as secondary sources. The main backbone of the search consisted of, but was not limited to the following keywords: “COVID-19 OR SARS-CoV-2 OR coronavirus”, “vaccine OR vaccination”, “multiple sclerosis OR MS”, and “disease modifying therapies OR disease modifying drugs OR DMT OR DMD”. As the COVID-19 vaccines were available after early 2021, the search was restricted to the studies after January 2021 and was conducted on November 7, 2021. Furthermore, a Google Scholar weekly alert was set, enabling us to screen the new results after the initial search date. The last screening of the new results was done in January 27, 2022.

After conducting the searches independently, two review authors used the Mendeley application for duplicate removal and screening of the titles and the abstracts of the results one by one. The possibly eligible studies were sought for retrieval of the full texts. Any errata or other linked citations were also retrieved. The reference lists of the possibly eligible studies were also scanned for more possibly eligible studies. All of the search results were archived in a reference manager file format, including a record of the excluded studies along with the reasons for exclusion.

#### 1.2. Eligibility

The exclusion of studies from syntheses was based on the following criteria and priorities:

1. Not a primary investigation;
2. Retracted/withdrawn;
3. No eligible participants;
4. No eligible exposures; and
5. No eligible comparators.

Nine group of DMTs were considered as eligible exposures and receiving no DMTs –either pwMS receiving no DMTs or healthy participants– was considered as the eligible comparators. For the further syntheses addressing the effects of dosing intervals among pwMS on BCDT, the fifth criterion was ignored.

The eligibility criteria for the participants were defined as:

1. No history/evidence of previous COVID-19; and
2. No history of corticosteroid administration within two months.

Three review authors independently assessed the full texts for eligibility. The papers that at least two review authors consider eligible were included in the study, and others were documented along with their reason for exclusion.

#### 1.3. Assessment of Risk of Bias

The National Institutes of Health Study Quality Assessment Tools were used by three review authors (raters) independently to assess the risk of bias in different levels of the studies. The source of each risk-of-bias judgment of the raters, along with detailed explanations and the final results of each assessment, is presented narratively and summarized in a separate ***Supplementary File***.

#### 1.4.  Data Extraction

Considering the heterogeneity of the used assays and their measuring units – which was anticipated – in order to present more practical/translational syntheses and facilitate the data extraction and synthesis processes, the data were extracted in a dichotomized fashion based on the seropositivity cut-off indices of the assays used in the studies. Hence, the number of seropositive and total participants were extracted, stratified by DMT exposure status, and with the unexposed (UX) people (healthy controls and/or pwMS on no DMTs) set as the comparator for all other DMTs. When piloting the data extraction process, we noticed that in most studies, the number of participants with negative post-vaccination serostatus would be zero for the UX cohorts and most DMT cohorts, i.e., the “zero” cells will cause computational problems for calculation of the effect measures. Hence, we decided to use the Peto method for estimation of odds ratio (OR) and 95% confidence interval (95%CI), which avoids the addition of a fixed continuity correction factor and has shown to feasibly provide unbiased estimations for relatively balanced cohorts ^1^. When neither the specific DMT nor the UX cohorts contained seronegative participants, no relative effect measurement was possible; therefore, those studies were only reported narratively. Additionally, due to the usage of the Peto method, in order to prevent biased estimations, we decided to exclude from synthesis and present narratively the measures calculated from studies with arms containing less than five total participants and/or considerably uneven arms.

Furthermore, for extraction of further measures pertaining to the dosing intervals of infused aCD20, unstandardized beta coefficient (B) along with 95%CI was calculated manually by implementing a univariate logistic regression model on the descriptive measures presented in the study.

Another issue identified when piloting the data extraction was that some studies assessed their specified outcomes in multiple time points, including before the first dose, before the second dose, and two to six weeks after the second dose. We only extracted the measures pertaining to the latest timepoints after the second dose, limiting the probability of missing the outcomes that took more time to occur. Data extraction piloting also revealed inconsistent reporting of the effect of baseline CD19/CD20-positive B-cell counts among pwMS on BCDT, preventing us from extracting and synthesizing them.

In case the studies used more than one assay for detecting humoral responses to SARS-CoV-2 – e.g., total anti-Spike (S) IgG, anti-S1 subunit IgG assays –including anti-receptor-binding domain (RBD) assays, anti-nucleocapsid (N) IgG, or other immunoglobulins than IgG, results were extracted regarding all assays, separately. The anti-N IgG-positive participants who received mRNA vaccines were excluded from synthesis as it indicated previous COVID-19 contraction.

#### 1.5. Syntheses

After assessing heterogeneity, the extracted results were pooled using the Peto fixed-effects model. The egger’s test for asymmetry of funnel plot was conducted to assess the small study effects and possible publication bias. When the adequate number of studies were available, we adopted an assay-specific fashion in the synthesis, i.e., measures of anti-S1 (including RBD) IgG, anti-S (including trimeric S), interferon-gamma release, CD4+ and CD8+ activation-induced marker (AIM), and multiplex polymerase chain reaction (M-PCR) assays were pooled and presented separately; otherwise, the T-cell assays were presented narratively, and the B-cell assays were pooled all together.

The results of pooled analyses were presented in an extended forest plot, funnel plots, and all outcomes which could not be entered into the pooled analysis were presented narratively. A meta-regression analysis was planned. However, it was not conducted due to several limitations, including our restrictions in obtaining data from the primary investigators. Hence, the possible reasons behind heterogeneity were only discussed narratively. We also found a few studies assessing the responses to boosters and narratively synthesized them, although this was not planned in our initial protocol.

Sensitivity analyses by performing all analyses on the subgroup of outcomes from the studies with “good” quality (least risk of bias) was done but was not presented as it did not show any change in any analysis, except for lower statistical power. A meta-analysis while adjusting the study weights based on the sensitivity of the used assays was initially planned; however, as all the studies used assays with more than 90% sensitivity, it was not conducted. The GRADE approach was used by two review authors to assess the certainty of the evidence. The baseline certainty of evidence was considered as moderate –due to observational nature of the synthesized studies, however, they were upgraded to high in case they were completely supported by non-dichotomized analysis in the individual studies. The judgments and arguments for down- or upgrading using GRADE was presented and justified to ensure transparency. The final assessment of certainty was presented in Table n (summary of findings), along with the number of included participants, effect measures and CI of the outcomes.

#### 1.6. Software

The Mendeley Desktop software version 1.19.8 for MacOS (Mendeley Ltd.) was used for management and screening of the studies. The Review Manager (RevMan) software version 5.4.1 for MacOS (The Cochrane Collaboration) was used for data extraction and synthesis. The SPSS software version 23 for MacOS (IBM Inc.) was used for the manual analyses.

#### 1.7. Registration and Approval

This study was registered in PROSPERO before initiation (id: CRD42021278107). No Institutional Review Board / ethics committee approvals were required for this study according to the national guidelines, as it did not involve human subjects.

### 2. Supplementary Figures

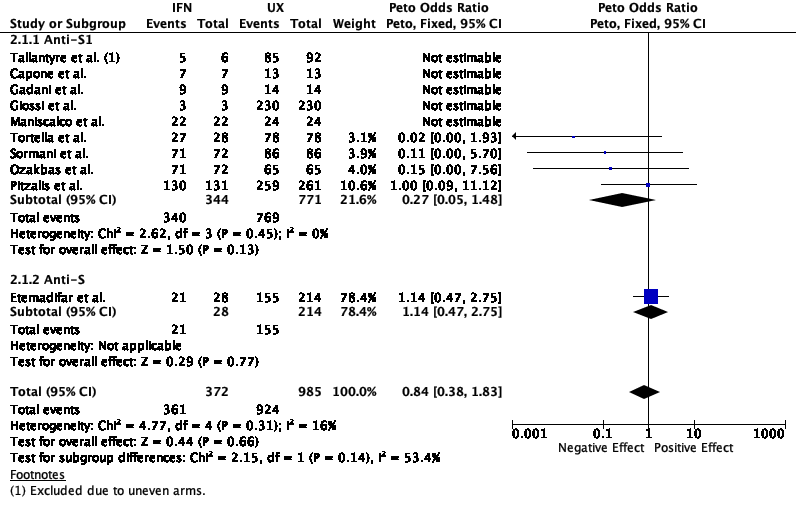

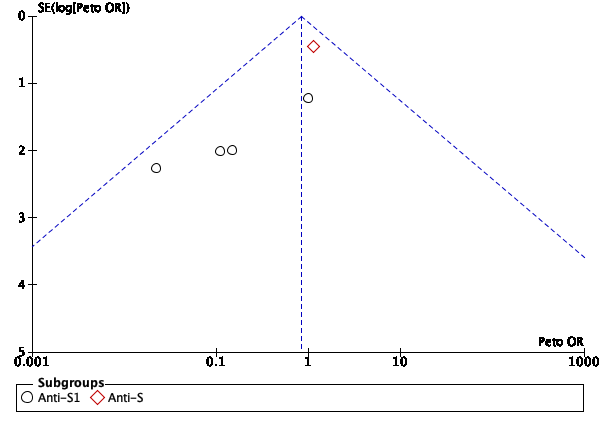

Supplementary Figure 1; Results of individual studies, heterogenicity tests, forest and funnel plots of studies measuring humoral response in pwMS on IFN

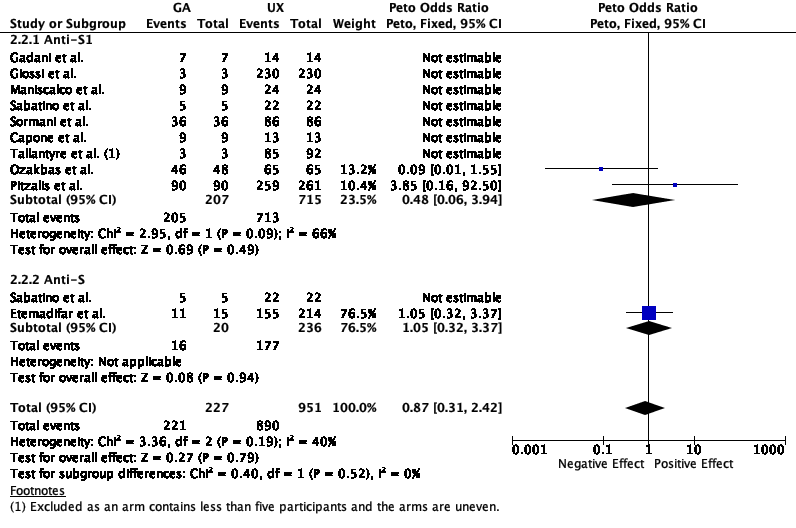

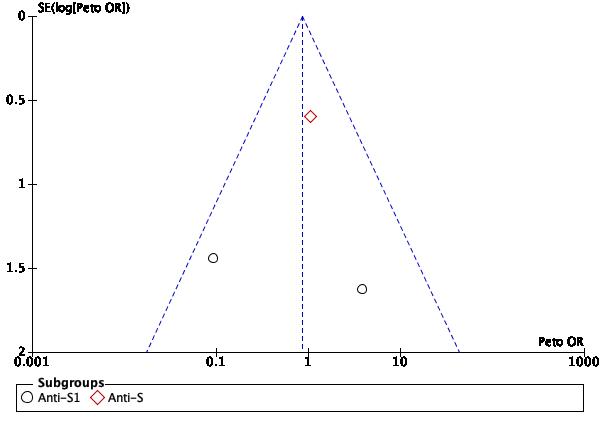

Supplementary Figure 2; Results of individual studies, heterogenicity tests, forest and funnel plots of studies measuring humoral response in pwMS on GA

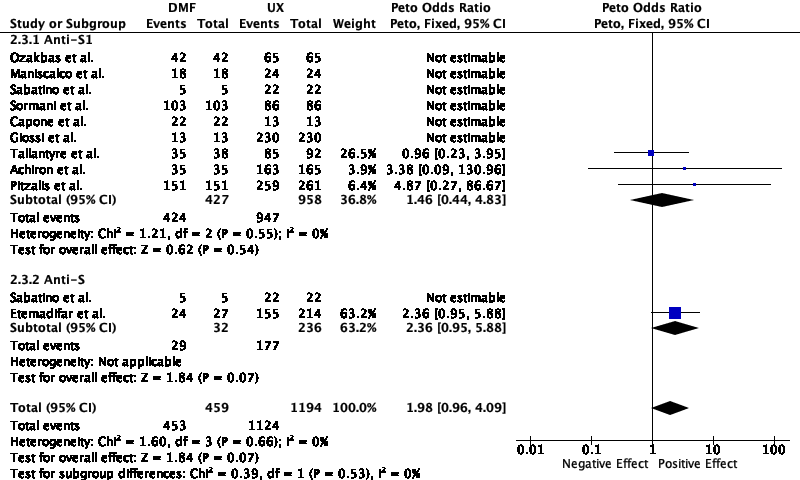

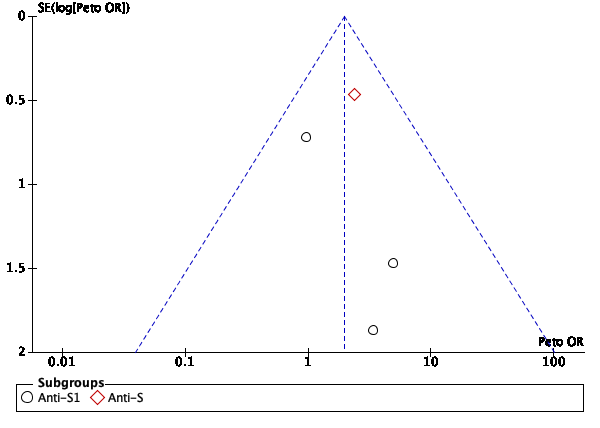

Supplementary Figure 3; Results of individual studies, heterogenicity tests, forest and funnel plots of studies measuring humoral response in pwMS on DMF

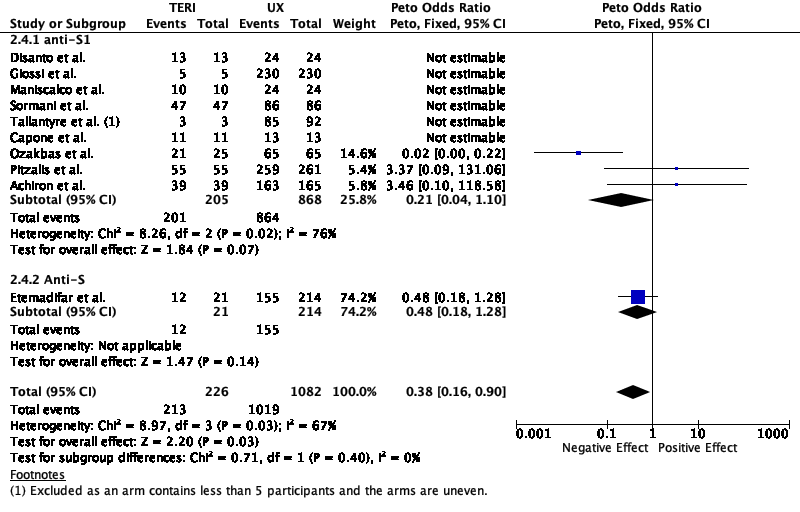

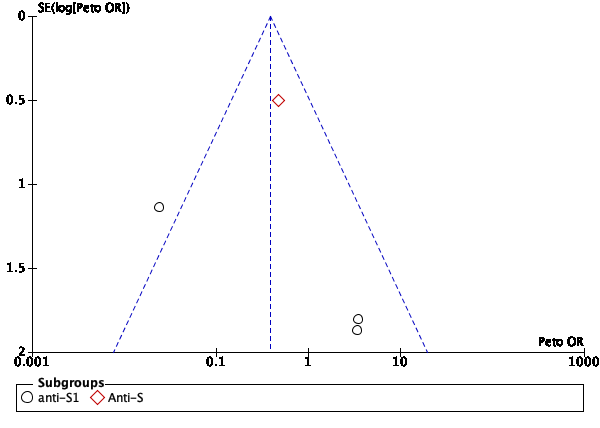

Supplementary Figure 4; Results of individual studies, heterogenicity tests, forest and funnel plots of studies measuring humoral response in pwMS on TERI

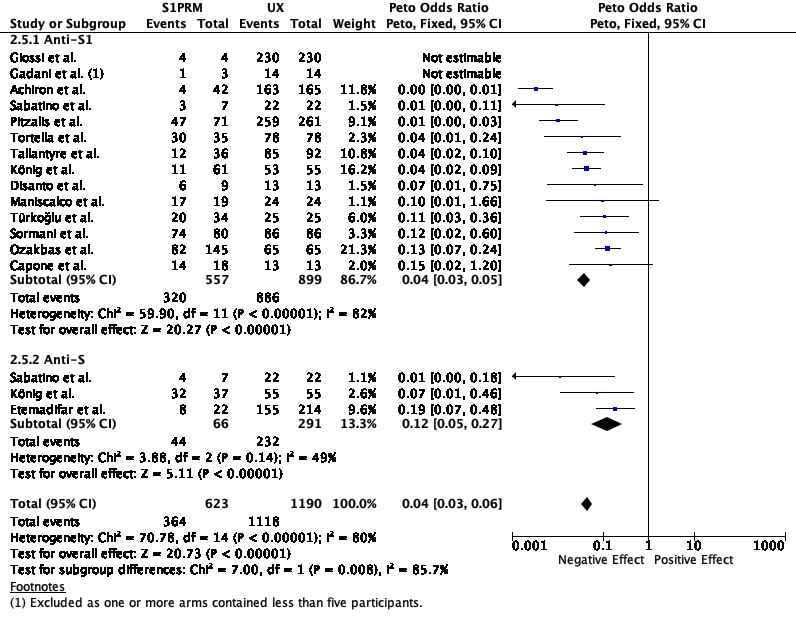

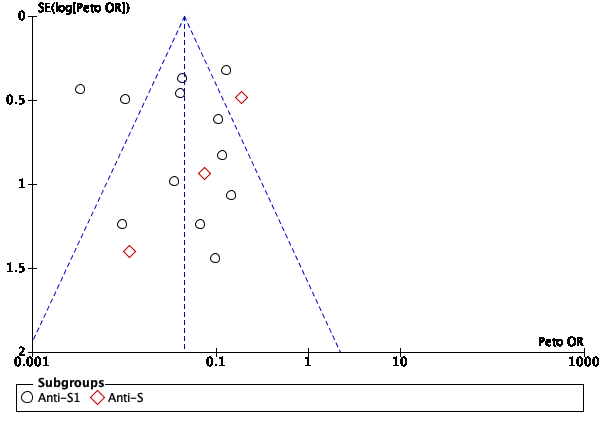

Supplementary Figure 5; Results of individual studies, heterogenicity tests, forest and funnel plots of studies measuring humoral response in pwMS on S1PRM

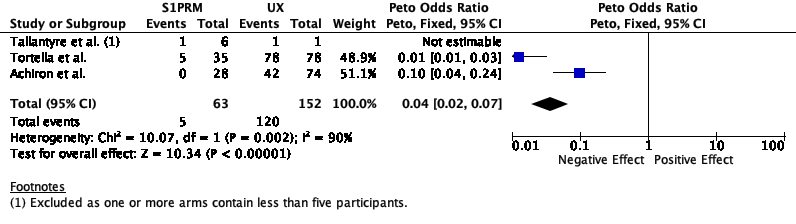

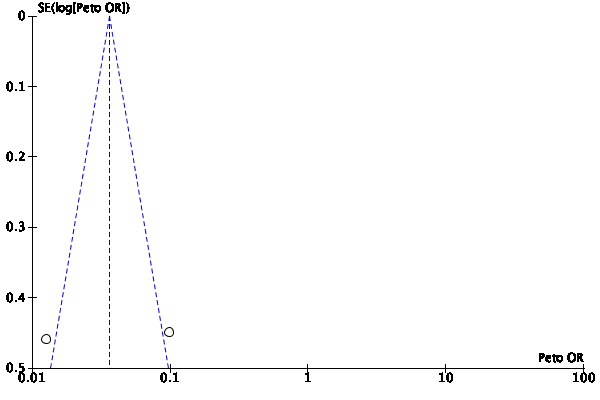

Supplementary Figure 6; Results of individual studies, heterogenicity tests, forest and funnel plots of studies measuring interferon-gamma release response in pwMS on S1PRM

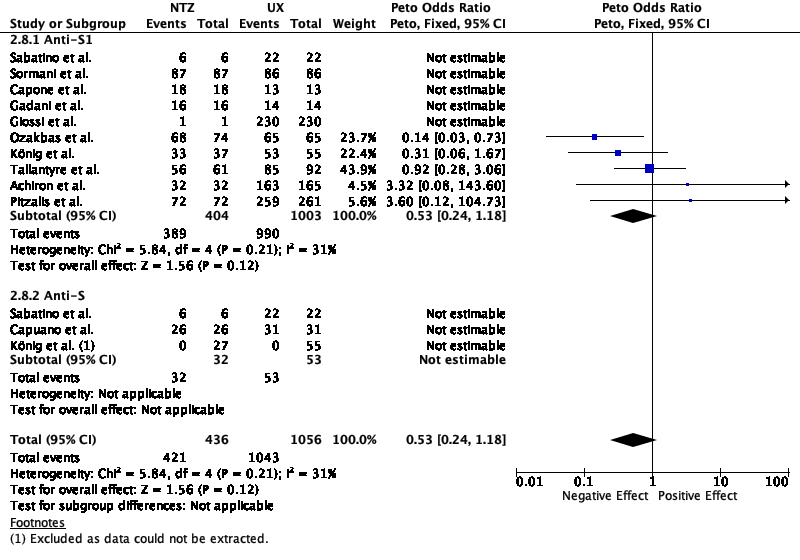

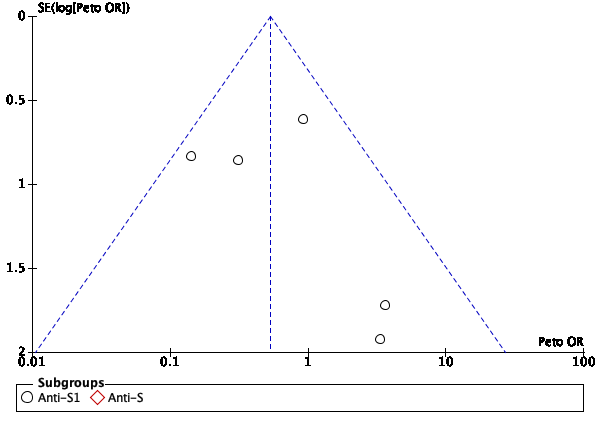

Supplementary Figure 7; Results of individual studies, heterogenicity tests, forest and funnel plots of studies measuring interferon-gamma release response in pwMS on NTZ

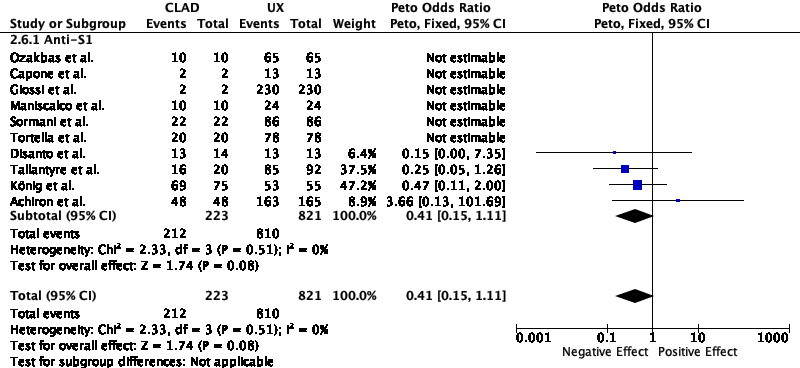

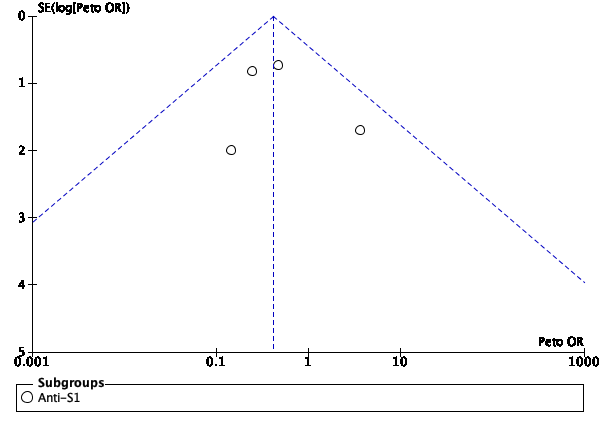

Supplementary Figure 8; Results of individual studies, heterogenicity tests, forest and funnel plots of studies measuring humoral response in pwMS on CLAD

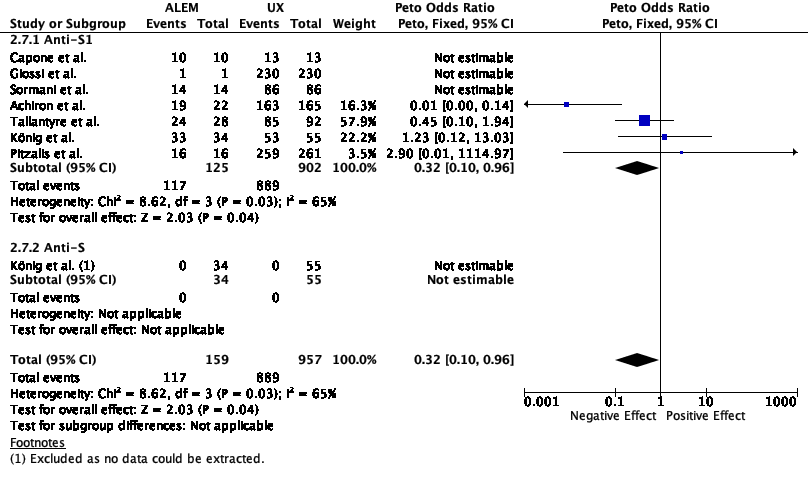

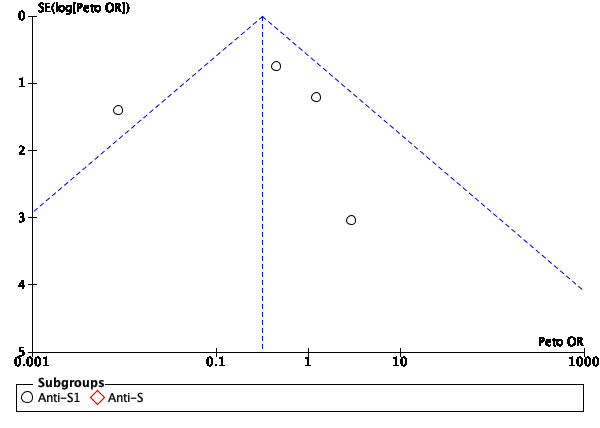

Supplementary Figure 9; Results of individual studies, heterogenicity tests, forest and funnel plots of studies measuring humoral response in pwMS on ALEM

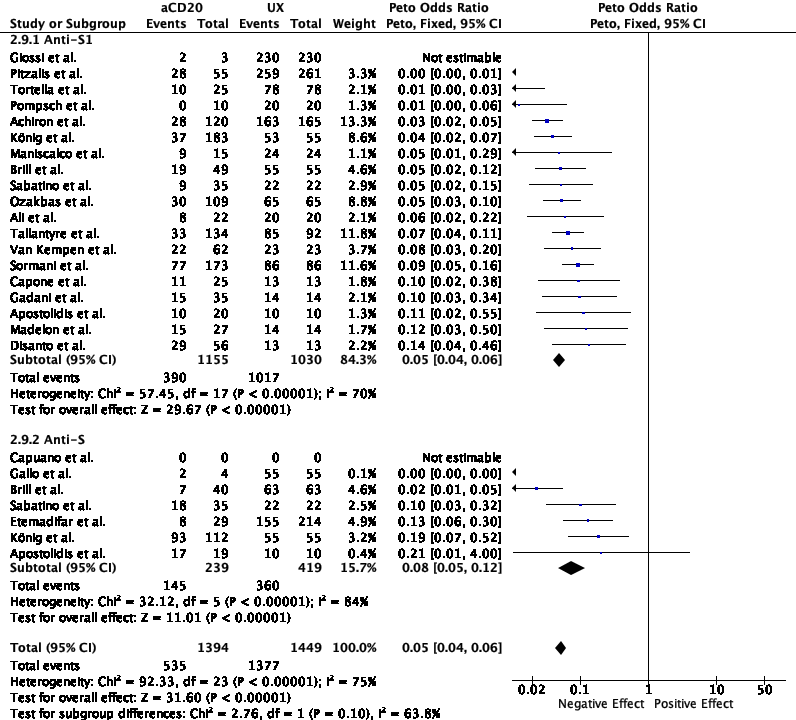

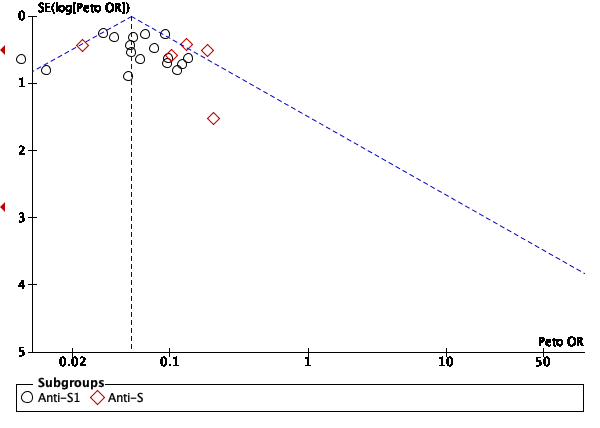

Supplementary Figure 10; Results of individual studies, heterogenicity tests, forest and funnel plots of studies measuring humoral response in pwMS on aCD20

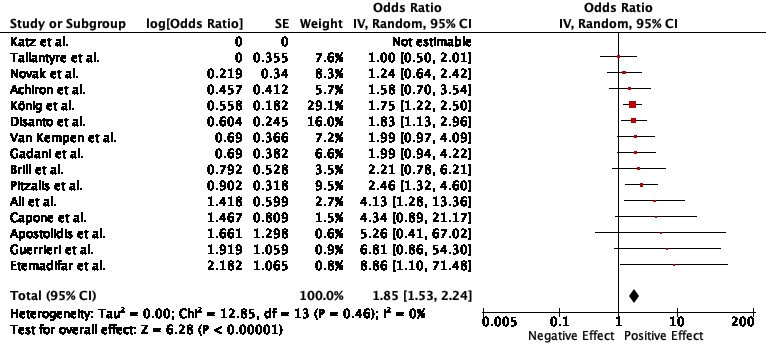

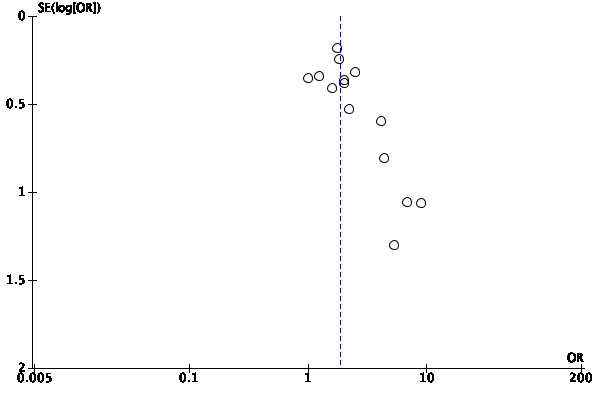

Supplementary Figure 11; Results of individual studies, heterogenicity tests, forest and funnel plots of studies measuring the effect of aCD20-to-vaccine period on humoral response

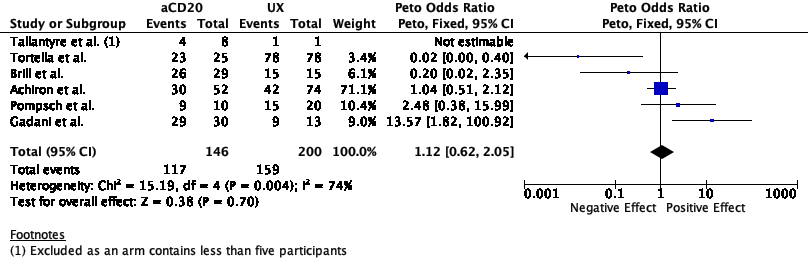

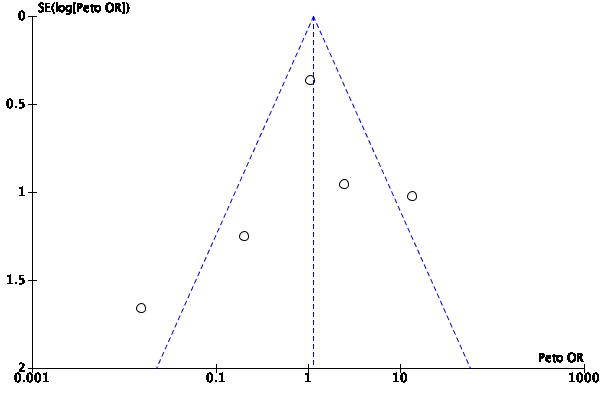

Supplementary Figure 12; Results of individual studies, heterogenicity tests, forest and funnel plots of studies measuring interferon-gamma release response in pwMS on aCD20

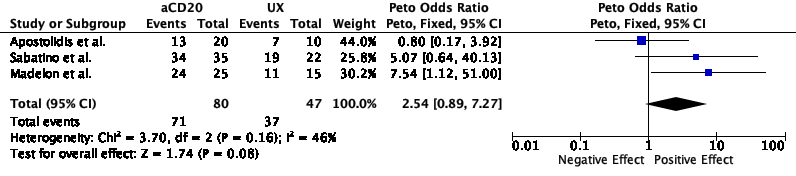

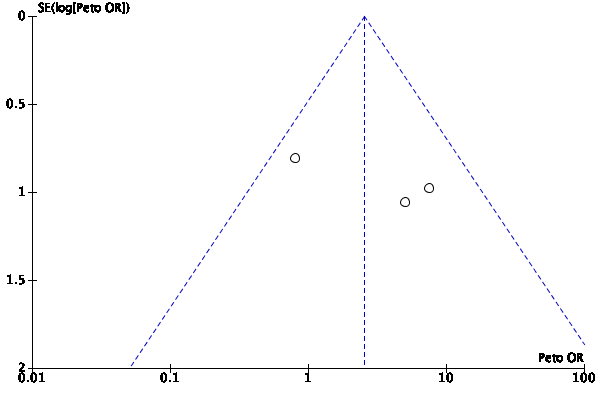

Supplementary Figure 13; Results of individual studies, heterogenicity tests, forest and funnel plots of studies measuring CD8+ AIM response in pwMS on aCD20
