## Supplementary material for "Multiple sclerosis disease-modifying therapies and COVID-19 vaccines: A practical review and meta-analysis": Quality Assessment

NIH Quality Assessment Tool for Observational Cohort and Cross-Sectional Studies available at: <https://www.nhlbi.nih.gov/health-topics/study-quality-assessment-tools>

| Study | Criteria | | | | | | | | | | | | | | Quality* |
| --- | --- | --- | --- | --- | --- | --- | --- | --- | --- | --- | --- | --- | --- | --- | --- |
|  | 1 | 2 | 3 | 4 | 5 | 6 | 7 | 8 | 9 | 10 | 11 | 12 | 13 | 14 |  |
| Achiron et al. | Y | Y | N/R | Y | NO | Y | Y | Y | Y | N/A | Y | N/R | Y | Y | Good |
| Capone et al. | Y | Y | Y | Y | NO | Y | Y | Y | Y | N/A | Y | N/R | Y | Y | Good |
| Capuano et al. | Y | NO | Y | Y | N/R | Y | Y | Y | Y | N/A | Y | N/R | Y | Y | Good |
| Disanto et al. | Y | Y | N/R | Y | NO | Y | Y | Y | Y | N/A | Y | N/R | Y | Y | Good |
| Etemadifar et al. | Y | Y | N/R | Y | Y | Y | Y | Y | Y | N/A | Y | N/R | N/R | Y | Good |
| Maniscalco et al. | Y | Y | Y | Y | NO | Y | Y | Y | Y | N/A | Y | N/R | Y | Y | Good |
| Ozakbas et al. | Y | Y | N/R | Y | NO | Y | Y | Y | Y | N/A | Y | N/R | Y | Y | Good |
| Sormani et al. | Y | Y | Y | Y | Y | Y | Y | Y | Y | N/A | Y | N/R | NO | Y | Good |
| Ali et al. | Y | Y | N/R | N/R | NO | Y | Y | Y | Y | N/A | Y | N/R | N/R | Y | Fair |
| Apostolidis et al. | Y | NO | N/R | N/R | N/R | Y | Y | Y | Y | N/A | Y | Y | N/R | N/R | Fair |
| Bigaut et al. | Y | Y | N/R | NO | NO | Y | Y | Y | Y | N/A | NO | N/R | N/R | Y | Fair |
| Brill et al. | Y | Y | N/R | NO | NO | Y | Y | Y | Y | N/A | Y | N/R | N/R | Y | Fair |
| Gadani et al. | Y | NO | N/R | Y | N/R | Y | Y | Y | Y | N/A | Y | N/R | Y | Y | Fair |
| Gallo et al. | Y | Y | N/R | Y | NO | Y | NO | Y | Y | N/A | Y | N/R | Y | N/R | Fair |
| Giossi et al. | Y | Y | N/R | Y | NO | Y | Y | Y | Y | N/A | Y | N/R | Y | N/R | Fair |
| Katz et al. | Y | Y | N/R | Y | NO | Y | NO | Y | Y | N/A | Y | N/R | Y | Y | Fair |
| König et al. | Y | Y | N/R | Y | NO | Y | Y | Y | Y | N/A | Y | N/R | Y | N/R | Fair |
| Madelon et al. | Y | NO | N/R | Y | NO | Y | Y | Y | Y | N/A | Y | N/R | Y | Y | Fair |
| Pitzalis et al. | Y | Y | N/R | NO | NO | Y | Y | Y | Y | N/A | Y | N/R | Y | Y | Fair |
| Pompsch et al. | Y | NO | N/R | Y | N/R | Y | Y | Y | Y | N/A | Y | N/R | N/R | Y | Fair |
| Sabatino et al. | Y | NO | N/R | Y | NO | Y | Y | Y | Y | N/A | Y | N/R | Y | N/R | Fair |
| Tallantyre et al. | Y | NO | N/R | Y | N/R | Y | Y | Y | Y | N/A | NO | N/R | Y | Y | Fair |
| Tortorella et al. | Y | NO | N/R | Y | NO | Y | Y | Y | Y | N/A | Y | N/R | Y | Y | Fair |
| Türkoglu et al. | Y | N | N/R | Y | NO | Y | Y | Y | Y | N/A | Y | N/R | Y | Y | Fair |
| VanKempen et al. | Y | NO | Y | Y | NO | Y | Y | Y | Y | N/A | Y | N/R | Y | N/R | Fair |

*Good: least risk of bias; Fair: low risk of bias; Poor: moderate/high risk of bias.

Study: Achiron et al.	DOI: 10.1016/j.jneuroim.2021.577746

| 1 | YES | The aim of study is clearly explained. |
| --- | --- | --- |
| 2 | YES | (who): patients with multiple sclerosis - (where): Sheba MS Center – (when): between December 2020 and February 2021 |
| 3 | N/R |  |
| 4 | YES | The inclusion and exclusion criteria were applied uniformly among all patients. |
| 5 | NO | No prior-to-study estimates on minimum required sample size are reported. |
| 6 | YES | This study was performed prospectively; exposure was assessed prior to outcome measurement. |
| 7 | YES | A minimum of interval between the second dose and serum sample acquisition of 28 days was set, which is long enough to detect the outcome. |
| 8 | YES | DMTs (exposure) were categorized in terms of medication type. Also, time from the last treatment dose and the first vaccination was taken into account and analysis of effect on outcome measure. |
| 9 | YES | The exposure measures in this study are parts of the patients’ medical history, which are usually objective and reliable if registered by health care providers. |
| 10 | N/A | The exposure measures in this research scenario do not require multiple assessments at different time points. |
| 11 | YES | The outcome measurement methods were clearly defined and implemented consistently across all study participants.  [Serum samples were examined for anti-SARS-COV-2 IgG using ELISA kit based on the recombinant S1 protein from the SARS-COV-2 spike protein (Euroimmun, Lubeck, Germany). Index values (signal to cut-off ratios) >1.1 were considered positive (EUROIMMUN. Anti-SARS-COV-2 ELISA IgG, [Package Insert], Moutain Lakes, NJ: EUROIMMUN US, 2020). Absolute lymphocyte count (ALC, cells/mm3) in the peripheral blood were collected at the same date of IgG serology and determined by a DxI hematology analyzer (Beckman Coulter USA).] |
| 12 | N/R |  |
| 13 | YES | No follow-up losses are reported. |
| 14 | YES | Possible covariates were taken into account in the analysis of treatment groups and healthy subjects. |

Study: Ali et al.	DOI: 10.1016/j.vaccine.2021.08.078

| 1 | YES | The probable factors impacting the outcome aimed in the study are documented. The necessity of performing the study is explained. The aim is clearly stated. | YES |
| --- | --- | --- | --- |
| 2 | YES | (who): participants with MS and other demyelinating diseases (Those who completed 2 doses of SARS-CoV-2 mRNA vaccines)- (where): University of Michigan Multiple Sclerosis Center- (when): between December 21, 2020 and May 19, 2021. | YES |
| 3 | N/R |  | N/R |
| 4 | NO | Patients were selected from one single center (University of Michigan Multiple Sclerosis Center) but from different populations (people with multiple sclerosis and other demyelinating diseases). The included participants were patients who had received 2 doses of SARS-CoV-2 mRNA vaccines during the same time period; inclusion criteria were applied uniformly (December 21, 2020 and May 19, 2021) | NO |
| 5 | N/R |  | NO |
| 6 | YES | The exposure status of participant prior to the study is clear (disease modifying therapies); participants were recruited based on their exposure. | YES |
| 7 | YES | The second blood sample was obtained approximately 3 weeks after the second dose of vaccination. This time frame is sufficiently enough. | YES |
| 8 | YES | The DMTs were categorized into different groups (B-cell depleting and non-B-cell depleting); further, both “the time between vaccination and the last dose of DMT” and “the duration of treatment with DMTs”, as two possible measures of exposure were taken into account. | YES |
| 9 | NO | The exposure measures mentioned above were clearly defined, and are parts of the patients’ medical history, which are usually objective and reliable if registered by health care providers. | YES |
| 10 | N/A | The exposure measures in this research scenario, i.e., DMT type, DMT receiving duration, and interval between the last DMT dose and vaccination, do not require multiple assessments at different time points. | N/A |
| 11 | YES | [The Roche anti-SARS-CoV-2 nucleocapsid assay works is an electrochemiluminescent immunoassay (ECLIA) which utilizes recombinant biotinylated and ruthenium-labeled nucleocapsid protein. The Siemens SARS-CoV-2 Spike RBD total antibody assay works as a chemiluminescent immunoassay (CLIA) by utilizing recombinant S1 subunit receptor-binding domain as a biotinylated and acridinium ester-conjugated antigen.] | YES |
| 12 | N/R |  | N/R |
| 13 | N/R |  | N/R |
| 14 | YES | Age, BMI, and total treatment duration didn’t differ between negative and positive antibody groups. | YES |

Study: Apostolidis et al.	DOI: 10.1038/s41591-021-01507-2

| 1 | YES | The research questions and objectives are clearly stated. | YES |
| --- | --- | --- | --- |
| 2 | NO | (who): MS patients treated with anti-CD20 monotherapy (n=20) + healthy controls (n=10). (when): December 2020 - April 2021; (where): not specified | NO |
| 3 | N/R |  | N/R |
| 4 | N/R | Although subjects in each group (i.e., MS and HC) seem to have fulfilled uniform inclusion criteria, which were specified before the study was begun, details of the inclusion criteria are not stated. | N/R |
| 5 | N/R |  | N/R |
| 6 | YES | The exposure status of participant prior to the study is clear (anti-CD20 monotherapy); participants were recruited based on their exposure. | YES |
| 7 | YES | The follow-up time was longer that one month which is fairly sufficient in this study. | YES |
| 8 | YES | The authors have taken into account the type of the anti-CD20 (i.e., ocrelizumab and rituximab), the number of previous cycles of anti-CD20, and the interval between the last cycle and vaccination; which enables fairly sufficient power for stratification of exposure risk. | YES |
| 9 | YES | The exposure measures mentioned above were clearly defined, and are parts of the patients’ medical history, which are usually objective and reliable if registered by health care providers. | YES |
| 10 | N/A | The exposure measures in this research scenario do not require multiple assessments at different time points. | N/A |
| 11 | YES | [Plasma samples were tested for SARS-CoV-2-specific antibody by ELISA59. The estimated sensitivity of the test is 100% (95% confidence interval (CI), 89.1 to 100.0%) and specificity is 98.9% (95% CI, 98.0 to 99.5%). Plasmids encoding the recombinant full-length spike protein and the RBD were provided by F. Krammer and purified by nickel-nitrilotriacetic acid resin (QIAGEN). Monoclonal antibody CR3022 was included on each plate to convert optical density values into relative antibody concentrations.] | YES |
| 12 | YES | [All experiments were conducted in blinded fashion with designated members of the clinical team (who were not part of running the assays) having access to the sample key until data were collected, at which point all researchers were unblinded for the analysis.] | YES |
| 13 | N/R |  | N/R |
| 14 | N/R |  | N/R |

Study: Bigaut et al.	DOI: 10.1016/j.neurol.2021.05.001

| 1 | YES | The aim of study was clearly defined which analyzing humoral response after COVID-19 vaccination and COVID-19 contracting in MS patients on DMTs. |
| --- | --- | --- |
| 2 | YES | (who): MS patients with serological results after COVID-19 vaccination and 61 MS patients with serological results after COVID-19_ (when): between January and April 2021_ (where): MS center of Strasbourg, France. |
| 3 | N/R |  |
| 4 | NO | The time period for all patients was the same but the inclusion criteria differs, one group is people with multiple sclerosis receiving COVID-19 vaccination and the other is people with multiple sclerosis who contracted COVID-19. |
| 5 | NO | Given the different types of medications, receivers of which were included in the study; and also given the inclusion of two types of immune triggers (i.e., vaccine and virus), the sample size may have been insufficient for subgroup analysis with regards to different medication types. This is stated as a limitation of the study by the very authors as well. |
| 6 | YES | The last dose of DMTs consumed by the patients is documented from their medical history and participant were exposed to the DMTs before the aimed outcome. |
| 7 | YES | The serology was assessed more than one month after COVID-19 and COVID-19 vaccination. |
| 8 | YES | The DMTs were categorized to Glatiramer acetate, Interferon b-1a, Teriflunomide, Dimethyl fumarate, Natalizumab, Mycophenolate mofetil, S1PR modulator, Anti-CD20 monoclonal antibody; each dose of DMT received by patients during the study, was documented. |
| 9 | YES | The exposure measures mentioned above were clearly defined, and are parts of the patients’ medical history, which are usually objective and reliable if registered by health care providers. |
| 10 | N/A | The exposure measures in this research scenario do not require multiple assessments at different time points. |
| 11 | NO | Two different anti-SARS-CoV-IgG assays were used; no further information is given in this regard, e.g., how many serum samples were assessed by each tests. |
| 12 | N/R |  |
| 13 | N/R |  |
| 14 | YES | The univariate analysis incorporated a linear regression model adjusted for possible confounder, such as age and sex. |

Study: Brill et al.	DOI: 10.1001/jamaneurol.2021.3599

| 1 | YES | The exposure, the aimed outcome and the participants relevant to the aim, are clearly defined. |
| --- | --- | --- |
| 2 | YES | (who): people with MS (pwMS)- (where): Hadassah Medical Center in Jerusalem, Israel – (when): between December 2020 and April 2021 |
| 3 | N/R | The number of screened patients and the final eligible participants is not reported. |
| 4 | NO | The inclusion and exclusion criteria were not clearly reported. |
| 5 | NO | No pre-enrollment estimates on the minimum sample size to detect a difference are provided; the study is extrapolating in nature and this is not a fatal flaw in this case. |
| 6 | YES | Participants were recruited considering their exposure of interest (Ocrelizumab) and were followed up prospectively to assess the aimed outcome for a specific timeframe after vaccination. |
| 7 | YES | Serum samples were taken 2-4 weeks after the second vaccine dose; the time frame is fairly sufficient. |
| 8 | YES | The duration of treatment is documented, but it is not clear if the whole treatment duration included only ocrelizumab therapy, and if not, for how long has ocrelizumab been given. However, it is interpretable from the results, and not from the methods section, that the time from the last ocrelizumab infusion and vaccination were investigated. |
| 9 | YES | The exposure measures mentioned above were clearly defined, and are parts of the patients’ medical history, which are usually objective and reliable if registered by health care providers. |
| 10 | N/A | The exposure measures in this research scenario do not require multiple assessments at different time points. |
| 11 | YES | Outcome measures (dependent variables) were clearly defined, valid, reliable and used uniformly within all the participants. [Serology response was measured using Liaison SARS-CoV-2 S1/S2 IgG (DiaSorin) and spike receptor-binding domain (RBD) Architect SARS-CoV-2 IgG II Quant assay (Abbott Diagnostics) with a positive response defined by IgG titer of 19 or more or 50 or more arbitrary units (AU) per mL, respectively. T-cell immune response to SARS-CoV-2 was assessed by detecting interferon γ using T-SPOT Discovery SARS-CoV-2 (Oxford Immunotec), a modified enzyme-linked immunospot technology, IVD CE–marked assay, using freshly isolated peripheral blood mononuclear cells.] |
| 12 | N/R |  |
| 13 | N/R |  |
| 14 | YES | It is interpretable from the results and the supplemental data that possible confounders were considered in the association/correlation analyses; e.g., the confounder “age” did show an impact on the outcome measure “antibody levels”. |

Study: Capuano et al.	DOI: 10.1007/s10072-021-05397-7

| 1 | YES | the exposure (Natalizumab), the aimed outcome (humoral response) and the participants (people with MS (pwMS) and healthy controls receiving BNT162b2 mRNA Covid-19 vaccine) are clearly defined. |
| --- | --- | --- |
| 2 | NO | The time frame of patient enrollment is not clear; (who): people with MS (pwMS) treated with Natalizumab and healthy controls. (where): the healthy controls were enrolled in the Clinic of University of Campania ‘Luigi Vanvitelli’ Naples, Italy. |
| 3 | YES | 31 people with MS (pwMS) were screened and 26 were eligible for the study (83.87%). 31 healthy controls were recruited from a large dataset of healthy controls enrolled in a surveillance program at the Clinic of University of Campania ‘Luigi Vanvitelli’ Naples, Italy. |
| 4 | YES | The exclusion and inclusion criteria were applied uniformly across both groups. |
| 5 | N/R |  |
| 6 | YES | The exposure in this study was being treated with Natalizumab; assessment of which was done before the study was initiated. |
| 7 | YES | Although 7 days after the second dose seems shorter than other similar studies, it maintains a more than a month interval from the first dose. Given the mechanism of action of Natalizumab, our interpretation is that a 5-week period after the first dose of vaccination is a reasonable interval for a non-B-cell depleted immune system to mount humoral immune responses against SARS-CoV-2. |
| 8 | YES | The time from the last Natalizumab infusion and vaccination was documented and taken into account. |
| 9 | YES | The exposure measures mentioned above were defined and investigated; these measures are parts of the patients’ medical history, which are usually objective and reliable if registered by health care providers. |
| 10 | N/A | The exposure measures in this research scenario do not require multiple assessments at different time points. |
| 11 | YES | Outcome measures (serum anti-SARS-CoV-IgG presence) were clearly defined, valid, reliable and used uniformly within all the participants. [Sera were tested at virology laboratory of our University Hospital, using the LIAISON® SARS-CoV-2 TrimericSIgG assay (DiaSorin-S.p.A.), for the detection of IgG antibodies to SARS-CoV-2 spike protein including neutralizing antibodies.] |
| 12 | N/R |  |
| 13 | YES | Follow-up data from all of the 26 patients with MS included were presented; thus, no follow-up losses are expected. |
| 14 | YES | Age and sex were matched between the people with multiple sclerosis (pwMS) and the healthy controls; possible confounding factors were taken into account through their analyses. |

Study: Disanto et al.	DOI: 10.1001/jamaneurol.2021.3609

| 1 | YES | The aim of the study is clearly defined as if to confirm the previous studies and investigate a new exposure- outcome assessing. The previous studies and the exposures and outcomes of interest are clearly cited. |
| --- | --- | --- |
| 2 | YES | (who): people with MS (pwMS) older than 18 years old. - (when): between February 25, 2021,and May 11, 2021 – (where): Neurocenter of the southern Switzerland |
| 3 | N/R |  |
| 4 | YES | The participants were recruited from the same population. The same inclusion and exclusion criteria were applied uniformly among all participants. |
| 5 | NO | No pre-enrollment estimates on the minimum sample size to detect a difference are provided; the study is extrapolating in nature and this is not a fatal flaw in this case. |
| 6 | YES | Since this study is a prospective observational cohort study, thus the exposure to DMTs in people with MS (pwMS) was assessed in the beginning of the study, and the participants were followed up and assessed for the aimed outcome. |
| 7 | YES | The time frame between the second vaccine dose and serological test (21-35 days) seems fairly sufficient for the desired outcome (seroconversion) to surface. |
| 8 | YES | The DMTs were subdivided into 9 main categories; time from last anti-CD20 infusion was also documented and taken into account. |
| 9 | YES | The exposure measures in this study are parts of the patients’ medical history, which are usually objective and reliable if registered by health care providers. |
| 10 | N/A | The exposure measures in this research scenario do not require multiple assessments at different time points. |
| 11 | YES | Outcome measures (dependent variables) were clearly defined, valid, reliable and used uniformly within all the participants. [Quantification of IgG against SARS-CoV-2 spike receptor binding domain was performed using a chemiluminescence microparticle immunoassay (Abbott; quantification limits, 21- 40000AU/mL; cutoff for seropositivity = 50AU/mL).4 CD19+ B cells were measured in the first serum sample obtaining using fluorescence-activated cell sorting.] |
| 12 | N/R |  |
| 13 | YES | Only 4 patients after testing positive within 2 weeks prior to first vaccine dose were excluded. (4/120 – 3.34%) |
| 14 | YES | Patients receiving medical treatments influencing response to vaccines other than MS DMTs were excluded. Sex and age were adjusted. |

Study: Etemadifar et al.	DOI: 10.1016/j.msard.2021.103417

| 1 | YES | The aim of the study is clearly defined. The exposure and desired outcome are noted. Exposure: disease-modifying therapies (DMTs) – outcome: humoral response to COVID-19 inactivated virus vaccination – population: people with MS |
| --- | --- | --- |
| 2 | YES | (who): exposed group: people with multiple sclerosis (pwMS) on DMTs, unexposed group: people having no history of immunosuppression and absence of any special condition. – (where): Isfahan-Iran. – (when): from August until October 2021. |
| 3 | N/R |  |
| 4 | YES | The participants were recruited from the same population. The inclusion and exclusion criteria for all the participants were applied uniformly. |
| 5 | YES | The minimum sample size justification and power description are provided. |
| 6 | YES | This study is a retrospective study, and the participants were included considering their status of exposure (disease-modifying therapies (DMTs)). Patients were on DMTs before they receive vaccination. |
| 7 | YES | It is interpreted from the results (table 1) that nearly all second dose-to-phlebotomy values from the included participants were more than 14 days, which seems fairly sufficient for mounting a humoral immune response. |
| 8 | YES | The DMTs as the exposures, were categorized into 6 groups. Time from the last anti-CD20 infusion-to-first vaccination is also documented and taken into account. |
| 9 | YES | The exposure measures in this study are parts of the patients’ medical history, which are usually objective and reliable if registered by health care providers; cases included in this study were patients from three private neurology clinics, thus, a reliably registered record is expected. |
| 10 | N/A | The exposure measures in this research scenario do not require multiple assessments at different time points. |
| 11 | YES | Quantification of the post-vaccination humoral responses among the participants was performed using an anti-Spike IgG enzyme-linked immunosorbent assay (ELISA) kit (Quanti SARS-CoV-2 Anti-Spike IgG, Pishtazteb Diagnostics, Iran). [The mentioned kit has a reported sensitivity and specificity of 98.16% and 99.01%, respectively, and an approved accuracy (Pishtazteb, 2021). The testing was carried out per manufacturer’s instructions (Pishtazteb, 2021) three times for each specimen, and the mean results were reported quantitatively in relative units (RU)/ml with a cut-off index (COI) value equal and above eight considered positive.] |
| 12 | N/R |  |
| 13 | N/R |  |
| 14 | YES | Identifying, controlling, and accounting for the possible confounders in the analysis were performed and the possible confounders were considered. Confounding variable controlling was performed using Multivariable logistic regression model. |

Study: Gadani et al.	DOI: 10.1016/j.ebiom.2021.103636

| 1 | YES | The aim of the study is clearly defined. The exposure (anti-CD20 therapy) and desired outcome (humoral and T cell immune responses to SARS-CoV-2) are noted. Population: people with multiple sclerosis. |
| --- | --- | --- |
| 2 | NO | (who): people with multiple sclerosis (pwMS) who had recently received the COVID-19 vaccine and were part of the COVID-RIMS study. – (where): Johns Hopkins MS center – (when): not specified. |
| 3 | N/R | 101 patients volunteered for this study; the participation rate remains unclear. |
| 4 | YES | All participants were recruited from the same population. Similar inclusion and exclusion criteria were applied uniformly to all participants. |
| 5 | N/R |  |
| 6 | YES | Patients were on DMTs before they receive vaccination. Also, the participants were recruited when they had been recently vaccinated. |
| 7 | YES | The phlebotomy was performed variably 4 to 8 (average 6.8) weeks after the terminal COVID-19 vaccination dose. This timeframe is relatively sufficient. |
| 8 | YES | The exposures were subdivided into 2 categories. Injectable therapy and Oral therapy. Time from last infusion of anti-CD20 therapies was also taken into consideration. |
| 9 | YES | The exposure measures in this study are parts of the patients’ medical history, which are usually objective and reliable if registered by health care providers |
| 10 | N/A | The exposure measures in this research scenario do not require multiple assessments at different time points. |
| 11 | YES | Same humoral and T-cell response assays were consistently utilized for all participants. The outcome measures (dependent variables) were clearly defined, valid, reliable. |
| 12 | N/R |  |
| 13 | YES | Data from all of the 101 patients with MS included were presented; thus, no follow-up losses are expected. |
| 14 | YES | Age, sex, and time from first dose of COVID-19 vaccination blood collection were adjusted using a logistic regression model. |

Study: Gallo et al.	DOI: 10.1007/s10072-021-05397-7

| 1 | YES | The aim of the study is clearly defined. The exposure (Ocrelizumab) and desired outcome (humoral response to SARS‑CoV‑2 mRNA vaccine) are noted. Population: patients with multiple sclerosis and healthy controls |
| --- | --- | --- |
| 2 | YES | (who): people with multiple sclerosis (pwMS) on Ocrelizumab and healthy subjects. – (where): Neurology clinic, University of Campania “Luigi Vanvitelli”, Naples, Italy. – (when): Since January 5th, 2021, |
| 3 | N/R |  |
| 4 | YES | The inclusion and exclusion criteria were applied uniformly to all participants. Apparently, all participants were included in the same time period. |
| 5 | NO | No pre-enrollment estimates on the minimum sample size to detect a difference are provided. Only 4 patients are included, but, the study is extrapolating in nature and this is not a fatal flaw in this case. |
| 6 | YES | The exposure in this study is being treated with Ocrelizumab in people with multiple sclerosis. These participants were enrolled based on their exposure to Ocrelizumab. All subjects were seronegative at the baseline; the exposure had taken place prior to the outcome. |
| 7 | NO | Only 7 days after the second dose of vaccination, the serum samples were obtained. Given that subjects on ocrelizumab may need more doses of the vaccine to increase their chance of seroconversion, the incorporated interval seems less sufficient, even though it still is a month and a week later than the first dose. |
| 8 | YES | The exposure (ocrelizumab) is investigated at different levels, i.e., type, duration, previous DMT, and time from the last infusion to the first vaccine dose. |
| 9 | YES | The exposure measures in this study are parts of the patients’ medical history, which are usually objective and reliable if registered by health care providers |
| 10 | N/A | The exposure measures in this research scenario do not require multiple assessments at different time points. |
| 11 | YES | The outcome measures (dependent variables) were clearly defined, valid, reliable. [Sera were stored at − 20 °C and tested using the LIAISON® SARS-CoV-2 TrimericS IgG assay (DiaSorin S.p.A.,Saluggia, Italy), an indirect chemiluminescence immunoassay (CLIA) technology for the detection of serum IgG antibodies to SARS-CoV-2 trimeric spike protein (anti-TSP IgG), including neutralizing antibodies . Performance and interpretation of results were done in accordance with the manufacturer’s instructions, and the IgG titers were expressed in Binding Antibody Units (BAU), an international standard unit, with 33.8 BAU/mL as cut-off value.] |
| 12 | N/R |  |
| 13 | YES | All 4 patients had completed the follow-up. |
| 14 | N/R |  |

Study: Katz et al.	DOI: 10.1016/j.msard.2021.103382

| 1 | YES | The aim of the study is clearly defined. The exposure (Ocrelizumab) and desired outcome (Humoral and T-Cell Responses to SARS-CoV-2 Vaccination) are noted. Population: people with multiple sclerosis. |
| --- | --- | --- |
| 2 | YES | (who): people with multiple sclerosis. – (where): Elliot Lewis Center for Multiple Sclerosis Care (MA, USA). – (who): between March 2, 2021, and July 1, 2021. |
| 3 | N/R |  |
| 4 | YES | All participants were enrolled from the same population. Inclusion and exclusion criteria for being in the study were pre-specified and applied uniformly to all participants. |
| 5 | NO | No pre-enrollment estimates on the minimum sample size to detect a difference are provided. |
| 6 | YES | The exposure was assessed prior to the outcome. The investigators screened participants for the presence of SARS-CoV-2 antibodies prior to vaccination, to ensure the antibodies (outcome) were result of vaccination and not the preexisting antibodies. This study is a prospective observational cohort study. |
| 7 | YES | Serology tests were done 3 to 4 weeks after final dose of vaccination. This timeframe is relatively sufficient. |
| 8 | YES | The exposure (DMT) was categorized and inspected regarding the type (i.e., Ocrelizumab or Natalizumab), number of previous cycles, time between the last infusion and first vaccination. |
| 9 | YES | The exposure measures in this study are parts of the patients’ medical history, which are usually objective and reliable if registered by health care providers. |
| 10 | N/A | The exposure measures in this research scenario do not require multiple assessments at different time points. |
| 11 | YES | The outcome measures (dependent variables) were clearly defined, valid, reliable. [The Roche Elecsys Anti-SARS-CoV-2 S immunoassay was performed per the manufacturer’s regulations using the cobas e602 analyzer to identify individuals with an antibody response to SARS-CoV-2. This was done prior to the first COVID-19 vaccination dose and 3-4 weeks following the final vaccination dose. The Adaptive Biotechnologies T-Detect COVID Test was performed per the manufacturer’s regulations using the Kingfisher Flex 711 system and the MagMax DNA Multi-sample Ultra 2.0 reagent kits to identify individuals with an adaptive T-cell immune response to SARS-CoV-2.] |
| 12 | N/R |  |
| 13 | YES | Given the inclusion of follow-up data from all 48 participants in the final analysis, no losses in follow-up are expected. |
| 14 | YES | Demographic and clinical factors including weight, BMI, age, race, baseline level of disability, MS disease subtype, duration of disease, duration of treatment, time from prior relapse, and smoking status were considered and assessed as potential confounders and predictors of vaccine response (aimed outcome). Patients with multiple comorbidities were excluded to avoid immuno-senescence and other health conditions as confounders. |

Study: König et al.	DOI: 10.1136/jnnp-2021-327612

| 1 | YES | The aim of the study is clearly defined. The exposure (Humoral immunity to SARS-CoV-2 mRNA vaccination) and aimed outcome (DMTs) are noted. Population: people with multiple sclerosis. |
| --- | --- | --- |
| 2 | YES | (who): people with multiple sclerosis (pwMS). – (when): all patients who donated a blood sample were reported by 30 June 2021. – (where): Norway (invitations were sent on a national level) |
| 3 | N/R |  |
| 4 | YES | The inclusion and exclusion criteria were applied uniformly to all participants. Apparently, all participants were included in the same time period. |
| 5 | NO | No pre-enrollment estimates on the minimum sample size to detect a difference are provided. |
| 6 | YES | The exposure was assessed prior to the study initiation and outcome measurement. |
| 7 | YES | Participants were assessed for the aimed outcome 3 to 12 weeks after full vaccination which is fairly sufficient. |
| 8 | YES | The exposure (DMT) is categorized and investigated at different levels, i.e., type, time from the last infusion to the first vaccine dose, etc. |
| 9 | YES | The information acquisition about the exposure history (i.e., DMTs) was done through digital questionnaire and national MS registry, which seems relatively reliable and valid. |
| 10 | N/A | The exposure measures in this research scenario do not require multiple assessments at different time points. |
| 11 | YES | The outcome measures (dependent variables) were clearly defined, valid, reliable. [Antibodies to full length Spike (HexaPro) from SARS-CoV-2 and the receptor-binding domain (RBD) were measured using a bead-based Flow cytometric assay13 in all included patients 3–12 weeks after full vaccination. Post-immunisation IgG titres were used as a correlate of protection,14 and reduced immunity was assumed in cases of IgG <70 arbitrary units (AU) corresponding to a level which was lower than found in 99% of all healthy vaccinated subjects.]. |
| 12 | N/R |  |
| 13 | YES | No follow-up losses are reported; also, none is expected, given the inclusion of data from all 1155 participants in the final analysis. |
| 14 | N/R |  |

Study: Madelon et al.	DOI: 10.1101/2021.07.21.21260928

| 1 | YES | The objective of the study is clearly defined. It is clearly mentioned that the study includes exposed and control participants. Exposure : anti-CD20 therapy – outcome: T cell responses 2 to mRNA-based COVID-19 vaccines |
| --- | --- | --- |
| 2 | NO | (who): people with multiple sclerosis, people with rheumatic diseases, healthy controls |
| 3 | N/R |  |
| 4 | YES | For enrolling the participants (3 groups), general and group-specific inclusion and exclusion criteria were applied uniformly among all participants. No specific information on study enrollment/conduction period is given, however, it is negligible and a short variance of participants’ vaccination and enrollment is expected |
| 5 | NO | No pre-enrollment estimates on the minimum sample size to detect a difference are provided. |
| 6 | YES | The study is done prospectively. The Exposure (DMTs) were measured prior to outcome (T cell responses). |
| 7 | YES | The immune responses were assessed 30 day after the second dose of vaccination which is sufficient. |
| 8 | YES | The exposure in the participants (being treated with DMTs) were subdivided into 5 categories. Glucocorticoid, Methotrexate, Leflunomide, Rituximab, Ocrelizumab. The timeframe between the last dose of medications (exposure) receiving and the vaccination, the medications (exposure) doses receiving by the participants (pwMS and people with rheumatic diseases) were documented. |
| 9 | YES | The exposure measures in this study are parts of the patients’ medical history, which are usually objective and reliable if registered by health care providers. |
| 10 | N/A | The exposure measures in this research scenario do not require multiple assessments at different time points. |
| 11 | YES | All antibody measurements were performed utilizing the Elecsys platform (Roche Diagnostics) and seroconversion was defined as > 0.8 IU/ml. |
| 12 | N/R |  |
| 13 | YES | No follow-up losses are reported; also, none is expected, given the inclusion of data from all 59 participants in the final analysis. |
| 14 | YES | Individuals in the control group were matched for age. |

Study: Pitzalis et al.	DOI: 10.1101/2021.09.26.21264067

| 1 | YES | The objective of the study is clearly defined. The exposure (disease-modifying therapies), participants (people with multiple sclerosis) , and the aimed outcome (humoral response to BNT162b2 vaccine) is clearly described. |
| --- | --- | --- |
| 2 | YES | (who): people with multiple sclerosis (pwMS) and healthy controls. – (where): MS clinical centers in Cagliari and Sassari in Sardinia (Italy). – (when): between April and June 2021. |
| 3 | N/R |  |
| 4 | NO | Although patients were recruited from a relatively homogenous population as controls; the inclusion and exclusion criteria were not clearly defined. |
| 5 | NO | No pre-enrollment estimates on the minimum sample size to detect a difference are provided. |
| 6 | YES | The measurement of exposure had preceded the outcome. |
| 7 | YES | The serum samples were obtained 30 days after the second dose of vaccination which is fairly sufficient. |
| 8 | YES | The type of the DMTs received by each patient is documented along with treatment duration and time between the last dose of treatment and inoculation. |
| 9 | YES | The exposure measures in this study are parts of the patients’ medical history, which are usually objective and reliable if registered by health care providers. |
| 10 | N/A | The exposure measures in this research scenario do not require multiple assessments at different time points. |
| 11 | YES | [Detection of anti‐SARS‐CoV‐2‐S and anti‐SARS‐CoV‐2‐N antibodies in serum samples was performed using the electrochemiluminescence immunoassays Elecsys® Anti-SARS-CoV-S and Elecsys® Anti-SARS-CoV-N (Roche) on the automated Cobas e-411 analyzer, according to the manufacturer's instructions.] |
| 12 | N/R |  |
| 13 | YES | No follow-up losses are reported; also, none is expected, given the inclusion of data from all 912 patients in the final analysis. |
| 14 | YES | The potential confounders were considered, including sex, age, EDDS, and smoking status. |

Study: Sabatino et al.	DOI: [10.1101/2021.09.10.21262933](https://dx.doi.org/10.1101%2F2021.09.10.21262933)

| 1 | YES | The objective of the study is clearly defined. The exposure (disease-modifying therapies), participants (people with multiple sclerosis), and the aimed outcome (SARS-CoV-2 vaccineinduced antibody and T cell immunity) is clearly described. |
| --- | --- | --- |
| 2 | NO | (who): people with multiple sclerosis and healthy controls. (where): five UK MS centres (Cardiff, Newport, Nottingham, Royal London Hospital (Barts Health NHS Trust) and Swansea) – (when): not specified |
| 3 | N/R |  |
| 4 | YES | Inclusion and exclusion criteria were uniformly applied. |
| 5 | NO | No pre-enrollment estimates on the minimum sample size to detect a difference are provided. |
| 6 | YES | The measurement of exposures preceded the outcome. |
| 7 | YES | Blood samples were obtained two weeks (for Comirnaty/BNT162b2 and mRNA-1273) or four weeks (for Ad26.COV2) after final dose of vaccination. Follow-up time is fairly sufficient. |
| 8 | YES | The exposure in the participants (being treated with DMTs) were subdivided into 6 categories. glatiramer acetate (GA), dimethyl fumarate (DMF), natalizumab (NTZ), sphingosine- 1-phosphate receptor modulator (S1P), rituximab (RTX), ocrelizumab (OCR); time from last infusion of anti-CD20 mAb was also documented. |
| 9 | YES | The exposure measures in this study are parts of the patients’ medical history, which are usually objective and reliable if registered by health care providers. |
| 10 | N/A | The exposure measures in this research scenario do not require multiple assessments at different time points. |
| 11 | YES | The outcome measures (dependent variables) were clearly defined, valid, reliable.  [Spectrally distinct Luminex beads were conjugated with trimeric spike protein (residues 1-1213), spike RBD (residues 328-533) (generously provided by Dr. John Pak, Chan Zuckerberg Biohub), or bovine-specific albumin fraction V (BSA) (Sigma-Aldrich #10735094001) at a concentration of 5 μg of protein per 1 million beads. Conjugation was done via an EDC/sulfo-NHS coupling strategy to terminal amines using antibody coupling kit following manufacturer’s instructions (Luminex #40-50016) as performed previously. Antibody analysis is performed by coronaphage VirScan.] |
| 12 | N/R |  |
| 13 | YES | No follow-up losses are reported; also, none is expected, given the inclusion of data from all 80 participants in the final analysis. |
| 14 | N/R |  |

Study: Sormani et al.	DOI: 10.1016/j.ebiom.2021.103581

| 1 | YES | The aim of this study is clearly defined. Exposure (disease modifying therapies), outcome (side effects and immunogenicity of SARS-CoV-2 mRNA vaccination), and the population (people with multiple sclerosis) are mentioned. |
| --- | --- | --- |
| 2 | YES | (who): people with multiple sclerosis (pwMS). – (where): 35 MS Italian MS centers. - (when): between March 4, 2021 and July 9, 2021. |
| 3 | YES | 85% of enrolled patients accepted to participate. |
| 4 | YES | Participants were recruited from a relatively homogenous population (MS centers across Italy) Inclusion and exclusion criteria were applied uniformly across all patients. |
| 5 | YES | The results in this article are interim results of an ongoing, large study; the planned number of participants is given, i.e., 2000. |
| 6 | YES | This study is performed prospectively and the exposures were measured prior to the outcome. |
| 7 | YES | The follow-up time of this study is relatively sufficient. |
| 8 | YES | The DMTs were categorized into 11 groups. Time since last infusion of an anti-CD20 agent (rituximab or ocrelizumab) is reported |
| 9 | YES | Data on patients’ exposure were presented by their neurologists, as part of their medical history, registered by the respective clinics; thus, the data acquisition method appears reliable. |
| 10 | N/A | The exposure measures in this research scenario do not require multiple assessments at different time points. |
| 11 | YES | High-affinity pan-Ig antibodies to SARS-CoV-2 were measured by a centralized laboratory with a double-antigen sandwich-based electrochemiluminescence immunoassay (ECLIA), using commercial kits (Elecsys, Roche Diagnostics Ltd, Switzerland). |
| 12 | N/R |  |
| 13 | NO | 24 % of the eligible included patients didn’t have the blood sample assessed. 780/1022 (76%) |
| 14 | YES | Age, sex, BMI, EDSS level, disease duration, presence of comorbidities, antibody levels in the pre-vaccination samples and vaccine type were adjusted among participants. |

Study: Tallantyre et al.	DOI: 10.1101/2021.07.31.21261326

| 1 | YES | The aim of this study is clearly defined. Exposure (disease modifying therapies), outcome (serological response to COVID-19 vaccination), and the population (large multi-center cohort) are mentioned. |
| --- | --- | --- |
| 2 | NO | (who): people with multiple sclerosis. – (where): five UK MS centers (Cardiff, Newport, Nottingham, Royal London Hospital (Barts Health NHS Trust) and Swansea) – (when): not reported. |
| 3 | N/R |  |
| 4 | YES | The inclusion and exclusion criteria were applied uniformly among all participants. |
| 5 | N/R |  |
| 6 | YES | The participants were exposed to the DMTs before the study. |
| 7 | YES | The follow-up time is around 4 weeks which is fairly adequate. |
| 8 | YES | Exposures (DMTs) were categorized to different groups. Details of DMT start dates, doses, total time on DMT, were documented from participants medical report. |
| 9 | YES | Details of DMT, e.g., start date, dose, total duration, were extracted from the patients’ official medical records, as researchers were given permission to access by patients. Thus, the data acquisition method appears sound. |
| 10 | N/A | The exposure measures in this research scenario do not require multiple assessments at different time points. |
| 11 | NO | Dried blood spots were analyzed in two different laboratories with two different techniques.  [In UHW (376), samples were analysed with the COVID-SeroKlir two-step enzyme-linked immunosorbent assay (ELISA) (Kantaro Biosciences, USA - supplied by EKF Diagnostics, UK).  At QMUL(37) , samples were analysed using the Globody technique.] |
| 12 | N/R |  |
| 13 | YES | Only one of the participants was excluded after initiation of the study. |
| 14 | YES | Logistic and linear regression models were used to account for the impact of possible confounders. |

Study: Giossi et al.	DOI: 10.1016/j.msard.2021.103415

| 1 | YES | The aim of this study is clearly defined. Exposure (disease modifying therapies or immunosuppressant), outcome (serological response to BNT162b2 COVID-19 vaccination), and the population (people with multiple sclerosis) are mentioned. |
| --- | --- | --- |
| 2 | YES | (who): consecutive healthcare workers with multiple sclerosis and controls. – (when): From February 2nd to April 2nd, 2021. – (where): three participating centers in Italy (Fondazione I.R.C.C.S. Istituto Neurologico Carlo Besta, Milano; I.R.C.C.S. Fondazione Mondino, Pavia; Azienda Ospedaliera Policlinico Umberto I, Roma) |
| 3 | N/R |  |
| 4 | YES | The study population was relatively homogenous; the inclusion and exclusion were applied uniformly for each group of participants. |
| 5 | NO | No pre-defined sample size estimates were provided. |
| 6 | YES | This study is performed prospectively. Participants were exposed to medications prior to receiving vaccination. |
| 7 | YES | The follow-up duration is nine weeks from the first vaccine dose which is fairly sufficient. |
| 8 | YES | The exposures were subdivided to 9 groups; further data on starting date, last administration date, etc. were also investigated. |
| 9 | YES | The exposure measures in this study are parts of the patients’ medical history, which are usually objective and reliable if registered by health care providers. |
| 10 | N/A | The exposure measures in this research scenario do not require multiple assessments at different time points. |
| 11 | YES | The outcome measures (dependent variables) were clearly defined, valid, reliable. [Samples were analyzed with SARS-CoV-2 IgG II Quant (Abbott) on the Architect instrument (Abbott) to quantitatively and qualitatively detect IgG antibodies against the Spike protein S1 receptor-binding domain (RBD).] |
| 12 | N/R |  |
| 13 | Y | No follow-up losses are reported; also, none is expected, given the inclusion of data from all 39 MS patients in the final analysis. |
| 14 | N/R |  |

Study: Pompsch et al.	DOI:	10.1186/s42466-021-00158-5

| 1 | YES | The aim of this study is clearly defined. Exposure (Ocrellizuamb), outcome (T-cell response to COVID-19 vaccination), and the population (people with multiple sclerosis) are mentioned. |
| --- | --- | --- |
| 2 | NO | (who): people with multiple sclerosis. – (where): Outpatient clinic for MS of the Alfried Krupp Hospital in Essen, Germany. – (when): not reported |
| 3 | N/R |  |
| 4 | YES | The inclusion and exclusion criteria were applied uniformly to all participants. |
| 5 | N/R |  |
| 6 | YES | Participants were exposed to Ocrelizumab(exposure) prior to vaccination and outcome measurement (T-cell response) |
| 7 | YES | The assays were performed about 28 days after the second dose of vaccination which is fairly sufficient. |
| 8 | YES | The level of exposure to Ocrelizumab is investigated as time from last infusion to first inoculation. |
| 9 | YES | The exposure measures in this study are parts of the patients’ medical history, which are usually objective and reliable if registered by health care providers. |
| 10 | N/A | The exposure measures in this research scenario do not require multiple assessments at different time points. |
| 11 | YES | The outcome measures (dependent variables) were clearly defined, valid, reliable. [To assess SARS-CoV-2-specific cellular immunity, ELISpot assays were performed, using peptide pools of the Spike (S) 1, the S1/S2, the membrane (M) and the nucleocapsid (NC) protein (Miltenyi Biotec, Bergisch Gladbach, Germany) and an S1 protein of SARS-CoV-2 (S Sino, Sino Biological, Wayne, PA, USA).  Antibodies were determined by a CE marked Anti-SARS-CoV-2 IgG semi-quantitative ELISA (Euroimmun, Lübeck, Germany).] |
| 12 | N/R |  |
| 13 | N/R |  |
| 14 | YES | Healthy controls were matched for confounders, such as age and sex. |

Study: Tortorella et al.	DOI: 10.1212/wnl.0000000000013108

| 1 | YES | The aim of this study is clearly defined. Exposure (disease modifying therapies), outcome (anti Region-Binding-Domain (RBD) neutralizing antibodies and Spike (S)-specific T-cell response to full COVID-19 vaccination), and the population (people with multiple sclerosis) are mentioned. |
| --- | --- | --- |
| 2 | NO | (who): health care workers and patients with multiple sclerosis. – (where): MS Centre of the Department of Neurosciences of San Camillo Forlanini Hospital (Rome, Italy). – INMI Lazzaro Spallanzani. – (when): not reported |
| 3 | N/R |  |
| 4 | YES | Inclusion and exclusion criteria were applied uniformly among each group. |
| 5 | NO | No pre-estimated minimum of sample size with respect to the statistical power needed to detect a potential difference was reported. |
| 6 | YES | This study was performed prospectively. |
| 7 | YES | Blood samples were collected 23 days after the second dose of vaccination. This time frame is more than 3 weeks which is fairly sufficient. |
| 8 | YES | The DMTs (exposure) were subdivided into different categories; other treatment characteristics, e.g., duration, were assessed. |
| 9 | YES | The exposure measures in this study are parts of the patients’ medical history, which are usually objective and reliable if registered by health care providers. |
| 10 | N/A | The exposure measures in this research scenario do not require multiple assessments at different time points. |
| 11 | YES | The outcome measures (dependent variables) were clearly defined, valid, reliable. [IFN-γ levels were quantified in the plasma samples using an automatic ELISA (ELLA, Protein Simple).  Humoral response to vaccination was assessed by quantifying the anti-Nucleoprotein IgG and the anti-RBD IgG (Architect® i2000sr Abbott Diagnostics, Chicago, IL, USA).] |
| 12 | N/R |  |
| 13 | YES | No follow-up losses are reported; also, none is expected, given the inclusion of data from all 108 MS patients in the final analysis. |
| 14 | YES | Multivariable adjustment for confounding factors was done. |

Study: van kempen et al.	DOI: 10.1016/j.msard.2021.103416

| 1 | YES | The exposure, and the aimed outcome is clearly defined. |
| --- | --- | --- |
| 2 | NO | (who): people with multiple sclerosis. – (where): Amsterdam MS Center. – (when): not reported. |
| 3 | YES | Only 2 out of the initial 89 enrolled patients were excluded from the study. |
| 4 | YES | All patients with multiple sclerosis were recruited from same population.  The inclusion and exclusion criteria were uniformly applied to each group. |
| 5 | NO | No pre-estimated minimum of sample size with respect to the statistical power needed to detect a potential difference was reported. |
| 6 | YES | This study is performed prospectively. The group of exposed patients were receiving Ocrelizumab prior to the outcome. |
| 7 | YES | The time of follow up is around 28 days after the second vaccine dose which is sufficient. |
| 8 | YES | The exposure (ocrelizumab) was further assessed with respect to time from last infusion to vaccination, number of previous infusions, etc. |
| 9 | YES | Clinical data were retrieved from patients’ medical files, which can be considered reliable and valid. |
| 10 | N/A | The exposure measures in this research scenario do not require multiple assessments at different time points. |
| 11 | YES | The outcome measures (dependent variables) were clearly defined, valid, reliable.  [SARS-CoV-2 antibodies against RBD were measured at Sanquin using an IgG specific ELISA (Steenhuis et al., 2021) Fresh whole blood was drawn at baseline (prior to vaccination) for measuring CD19+ B-cells with a highly sensitive  assay (Koutsakos et al., 2021). a qualitative anti-RBD bridging assay was also used as this assay has better sensitivity to detect low levels of antibodies (Vogelzang et al., 2020).]. |
| 12 | N/R |  |
| 13 | YES | No follow-up losses are reported; also, none is expected, given the inclusion of data from all 87 MS patients in the final analysis. |
| 14 | N/R |  |

Study: Capone et al.	DOI: 10.1007/s13311-021-01165-9

| 1 | YES | The aim of study is clearly explained. |
| --- | --- | --- |
| 2 | YES | (who): people with multiple sclerosis. – (where): three MS centers of Rome, Italy (Università Campus Bio-Medico, Fondazione Policlinico Universitario “A. Gemelli” IRCCS, and San Filippo Neri Hospital). – (when): between 26 April 2021 and 4 June 2021. |
| 3 | YES | Only four out of 140 patients, initially enrolled, were excluded (previous COVID-19 infection). |
| 4 | YES | The inclusion and exclusion criteria were applied uniformly among all patients. |
| 5 | NO | No prior-to-study estimates on minimum required sample size were given. |
| 6 | YES | This study was performed prospectively. |
| 7 | YES | The blood test was performed 30 days after the second dose of vaccination. This time frame is sufficient. |
| 8 | YES | The DMTs (exposure) were subdivided into different categories. |
| 9 | YES | Clinical data were retrieved from patients’ electronic health records in MS centers, which can be considered reliable and valid. |
| 10 | N/A | The exposure measures in this research scenario do not require multiple assessments at different time points. |
| 11 | YES | The outcome measures were clearly defined, valid, reliable, and implemented consistently across all study participants.  [The detection of SARS-CoV-2 IgG antibodies in blood samples was performed using a chemiluminescent microparticle immunoassay for quantitative and qualitative detection of IgG against SARS-CoV-2 nucleoprotein (Spike-RBD S1) (SARSCoV- 2 IgG II for use with ARCHITECT; Abbott Laboratories, Abbott Park, IL, USA; ref: 6S60-22).] |
| 12 | N/R |  |
| 13 | YES | No follow-up losses are reported; also, none is expected, given that the whole study follow-up period was 1 month after second inoculation, and authors have reported that no case of SARS-CoV-2 infection has been detected among the participants during this period. |
| 14 | YES | Demographic and clinical factors of potential confounding effect were assessed and were not different between participants with different outcomes. |

Study: Maniscalco et al.	DOI: 10.1007/s13311-021-01165-9

| 1 | YES | The aim of study is clearly explained. |
| --- | --- | --- |
| 2 | YES | (who): people with multiple sclerosis. – (where): MS center of the Cardelli hospital, Naples, Italy. – (when): from March 2021 to June 2021. |
| 3 | YES | 85.1% (n=149) of eligible individuals (mentioned above) met the inclusion criteria. |
| 4 | YES | The inclusion and exclusion criteria were applied uniformly among all patients. All participants were Caucasian. |
| 5 | NO | No prior-to-study estimates on minimum required sample size are reported. |
| 6 | YES | This study was performed prospectively; exposure was assessed prior to outcome measurement. |
| 7 | YES | SARS-CoV-2 IgG was tested for 21 days after the second vaccination, which is relatively enough. |
| 8 | YES | The DMTs (exposure) were subdivided into different categories; in the case of depletant medications, interval between last dose and vaccination was also assessed. |
| 9 | YES | The exposure measures in this study are parts of the patients’ medical history, which are usually objective and reliable if registered by health care providers. |
| 10 | N/A | The exposure measures in this research scenario do not require multiple assessments at different time points. |
| 11 | YES | The outcome measurement methods were clearly defined, valid, reliable, and implemented consistently across all study participants.  [Quantitative determination of antibodies to the SARS-CoV-2 spike protein was carried out by Roche Elecsys® Anti‑SARS‑CoV‑2 S assay (Roche Diagnostics International Ltd, Rotkreuz, Switzerland). The assay was performed using a recombinant protein representing the RBD of the S antigen leading to a double-antigen sandwich assay complex which favors detection of high affinity antibodies against SARS-CoV-2 (range between 0.4 to 250 U/mL), resulting in a sensitivity of 98.8% (95% CI: 98.1 – 99.3%). |
| 12 | N/R |  |
| 13 | YES | No follow-up losses are reported; also, none is expected, given the inclusion of data from all 125 MS patients in the final analysis. |
| 14 | YES | Healthy controls were matched for age, and sex; possible confounding factors were adjusted for in the multilinear regression model. |

Study: Türkoglu et al.	DOI:10.1016/j.msard.2022.103524

| 1 | YES | The aim of study is clearly explained. |
| --- | --- | --- |
| 2 | NO | (who): consecutive patients with multiple sclerosis - (where): Amsterdam MS Center. – (when): not reported. |
| 3 | N/R |  |
| 4 | YES | The inclusion and exclusion criteria were applied uniformly among all patients. |
| 5 | NO | No prior-to-study estimates on minimum required sample size are reported. |
| 6 | YES | This study was performed prospectively; exposure was assessed prior to outcome measurement. |
| 7 | YES | SARS-CoV-2 IgG was tested for 28 days after each vaccination dose, which is relatively enough. |
| 8 | YES | The duration of therapy with DMTs (exposure) were measured and taken into account. |
| 9 | YES | The exposure measures in this study are parts of the patients’ medical history, which are usually objective and reliable if registered by health care providers. |
| 10 | N/A | The exposure measures in this research scenario do not require multiple assessments at different time points. |
| 11 | YES | The outcome measurement methods were clearly defined, valid, reliable, and implemented consistently across all study participants.  [Sera were collected 28 days after both first (day 28) and second (day 58) vaccinations and kept at 80 ◦C freezer until analysis. Immunoassay for the detection of SARS-CoV-2 IgG antibodies in sera was performed using Euroimmun (Luebeck, Germany) quantitative ELISA kit, designed for detection of antibodies to spike protein of the SARS-CoV-2 virus. The assay was performed following the manufacturer’s instructions and an index value higher than 1.1 was considered positive.] |
| 12 | N/R |  |
| 13 | YES | No follow-up losses are reported. |
| 14 | YES | Healthy controls were matched for age, and sex; no significant correlation was found between the outcome and possible confounding factors in the regression model. |

Study: Ozakbas et al.	DOI: 10.1016/j.msard.2022.103486

| 1 | YES | The aim of study is clearly explained. |
| --- | --- | --- |
| 2 | YES | (who): patients with multiple sclerosis - (where): Dokuz Eylul University Izmir, Turkey – (when): May 2021 and September 2021 |
| 3 | N/R |  |
| 4 | YES | The inclusion and exclusion criteria were applied uniformly among all patients. |
| 5 | NO | No prior-to-study estimates on minimum required sample size are reported. |
| 6 | YES | This study was performed prospectively; exposure was assessed prior to outcome measurement. |
| 7 | YES | A minimum of interval between the second dose and serum sampling of two-weeks was set, which appears relatively enough. |
| 8 | YES | DMTs (exposure) were categorized in terms of medication type. However, treatment duration, or time from last dose of treatment to vaccination/serum sampling was not studied in depth for a better quantification of exposure into a continuous measure. |
| 9 | YES | The exposure measures in this study are parts of the patients’ medical history, which are usually objective and reliable if registered by health care providers. |
| 10 | N/A | The exposure measures in this research scenario do not require multiple assessments at different time points. |
| 11 | YES | The outcome measurement methods were clearly defined and implemented consistently across all study participants.  [The primary outcome was the quantification of antibody response after two doses of the SARS-CoV-2 vaccine. Antibody levels were transformed on a Log10 scale to normalize their distribution, and the ‘AU/mL Log’ name was used after that. For antibody titer less than the detectable limit of 21AU/mL, in order to prevent missing data during Log10 transformation, a titer of 0.01Au/mL was used. Antibody titer of patients with MS who did not use DMT (pwMSw/oT) was compared as a separate group with pwMS with treatment and HC.] |
| 12 | N/R |  |
| 13 | YES | Only three (out of 593) patients were excluded from the study; one due to unsuccessful serum sample acquisition and the other due to incident hemolysis in the sample, and the other due to their medication type (RTX). |
| 14 | YES | Healthy controls were matched for age and sex. |
